# Economic evaluation of caregiver interventions for children with developmental disabilities: a scoping review

**DOI:** 10.1101/2024.10.23.24315995

**Authors:** Angela Kairu, Edwin Dzoro, Vibian Angwenyi, Charles Newton, Charlotte Hanlon, Rosa A Hoekstra, Amina Abubakar, Edwine Barasa

**Author notes:** Authors’ contributions: AK, CN, AA and EB conceptualized the study. Data was collected by AK and ED. AK and ED conducted the analysis. AK drafted the initial manuscript which was subsequently revised for important intellectual content by all authors. All authors read and approved the final manuscript.

## Abstract

**Introduction:** Globally, families with children with developmental disabilities (DDs) may experience several challenges, including social isolation, stigma, and poverty especially in low-income settings in Africa. Most children with DDs in Africa remain unidentified and receive no formal support. Caregiver interventions focusing on education and training for carers of children with DDs have been shown to be adaptable and low intensity in implementation. However, economic evaluation evidence on caregiver interventions for DD, which is important for effective resource allocation, is limited. This review aimed to describe the nature of evidence available and methodological aspects of economic evaluations for caregiver interventions for DDs.

**Methods:** This scoping review employed the Arksey and O’Malley framework and aligned with the Preferred Reporting Items for Systematic Reviews and Meta-Analyses extension for Scoping Reviews (PRISMA-ScR). Seven electronic databases, grey literature and cited references were systematically searched to identify eligible studies on economic evaluations of caregiver interventions for children with DDs published in 1993-2023. We assessed the quality of the included studies using the Drummond checklist. Data were systematically extracted, tabulated, and qualitatively synthesised using inductive thematic analysis.

**Results:** The searches yielded 7811 articles. Seventeen studies all in high-income countries met the inclusion criteria which focused on caregiver interventions for autism spectrum disorder (n=7), attention deficit hyperactivity disorder (ADHD) (n=6), disruptive behaviour and behaviour problems with ADHD (n=5), intellectual disabilities (n=1) and language delay (n=1).

The most used economic evaluation approach was trial based models (n=14), followed by decision analytic models (n=5)). The methods were not explicitly stated in 1 study. Economic evaluation analyses included cost effectiveness (n=11), costing (n=3), cost utility (n=2), cost consequence (n=1) cost benefit (n=1), and combined analyses (n=2). Nine studies reported the interventions as cost effective, five studies reported the intervention to be cost saving, and one identified caregiver costs as a cost driver. The main identified methodological challenges were related to costing, outcome measurement in children and the appropriate time horizon for modelling.

**Conclusion:** Caregiver interventions demonstrate cost-effectiveness, with the available evidence supporting the adoption of the interventions evaluated. Caregiver interventions are a promising avenue to strengthen access and reduce costs associated with health services for children with DDs. Additionally, this review identified key methodological challenges and highlighted areas for further research to address these limitations. Prioritizing more economic evaluation studies in this area would inform decision-making on efficient resource allocation, promote inclusivity and equitable access to services for children with DDs.

## Introduction

Developmental disabilities (DDs) are a heterogenous group of conditions characterized by impairments that affect a child’s physical, learning, or behavioural functioning (1). Globally, 53 million children under the age of 5 years have DDs such as sensory impairment, intellectual disability, epilepsy, and autism spectrum disorder (ASD) (2). With the lack of improvement of the global burden of developmental disabilities over the last three decades (2), the socioemotional consequences, academic challenges and economic cost to society will be sustained or may worsen over time (3). More specific to DDs, neurodevelopmental disorders (NDDs) occur in the developmental period and induce deficits that produce impairments of functioning (personal, academic, occupational or social) (4). They include intellectual disability (ID); ASD; Attention-Deficit/Hyperactivity Disorder (ADHD); communication disorders; neurodevelopmental motor disorders; and specific learning disorders (4). The prevalence of NDDs have rarely been assessed as a whole, with existing literature highlighting multimorbidity as a norm (3). The mmajority of children with DDs lack access to care especially in Africa, due to inadequate capacity of skilled human resource (5, 6). With focus on NDDs, substantial evidence shows that parents can learn skills to effectively improve their child’s development and positive behaviour (7).

Caregiver-focused interventions have been developed to train and educate parents/carers of children with developmental disabilities to support them in their child’s development (8). Caregiver interventions are often centred on increasing parental responsiveness to improve the child’s behaviour and communication outcomes (9–12). Parent skills training models have shown to be low-intensity interventions due to less practitioner time requirement, and the narrower focus of parenting strategies, and are adaptable for clinic, home, groups or individuals (13, 14). Further, interventions in early childhood are shown to have the strongest impact on improving the child’s long-term outcomes and parent outcomes (10–12). Therefore, the need to explore the childhood specific strategies is important to reduce the overall economic burden of the illness. As such, the use of economic evaluations on mental health interventions and strategies has steadily gained interest. The economic evaluation data provides policy makers with information on the best value for money for the interventions (15). Within resource constraints, it is important that health interventions are effective in reducing the burden of disease. Economic evaluations can provide this information, and it is important to assess the current scope of evaluations of child specific interventions to inform future policy decisions.

Economic evaluation is an approach that contributes evidence on the economic costs and outcomes of health interventions to inform efficient resource allocation within a budget constraint. (16). Policy strategies to decrease the impact of mental disorders are optimally effective when they are informed by evidence that determines how to improve efficacy, efficiency and cost-effectiveness of interventions, and their translation into clinical practice (17). With a goal of maximizing health and well-being, economic analyses are needed alongside effectiveness evidence for policy makers to identify the best options for mental health resources (18). Costs and cost-effectiveness data for caregiver interventions for children with DDs is scarce. Lamsal et al (2017) highlight few economic evaluations have been conducted to value interventions for children with NDDs resulting from challenges in applying economic evaluation methodologies for this heterogenous population (1). Further a review by Kularatna et al (2022) identified twelve model based economic evaluations for care for NDDs, however none modelled the impact on families and caregiver (19).

For childhood intervention programmes, some of the methodological challenges are related to cost and outcome measurement and valuation, the requirements for sensitivity analyses, the decision rules adopted by decision makers and the interpretation of results considering contextual factors (20, 21). Given these factors, there is a notable gap in literature that provides an overview of economic evaluations for caregiver interventions for children with DDs. The aim of this scoping review is to map the body of literature and describe the nature of evidence available on the economic evaluations for caregiver interventions for NDDs. Additionally, the review appraised the quality of studies included and discussed their methodological challenges and ways to mitigate them. By outlining the available evidence, we aim to inform the paediatric economic evaluation methodological approaches utilized and associated challenges and build on existing literature on interventions for child and adolescent mental healthcare.

## Methods

We applied the scoping review approach proposed by Arksey and O’Malley (22), and adhered to the Preferred Reporting Items for Systematic Reviews and Meta-Analyses extension for Scoping Reviews (PRISMA-ScR) guidelines for reporting this review. The review protocol was uploaded to Open Science Framework (OSF) https://osf.io/ymwr2.

### Search strategy and selection criteria

An extensive literature search was conducted using electronic databases that included PubMed, PsycINFO, Web of Science, the International Network of Agencies for Health Technology Assessment (INAHTA), Paediatric Economic Database Evaluation (PEDE), CINAHL, Econ Lit through EBSCO from January 1993 to December 2023. Further, we searched databases of grey literature specifically google scholar, and references of the included studies. The final search was conducted on September 18^th^, 2024. The search terms used in all search strategies were categorized into 4 blocks including: i) caregiver (parent, family); ii) interventions (e.g. training, programme, groups); iii) neurodevelopmental disorders (e.g. autism, intellectual disability); and iv) economic evaluation (e.g. cost utility analysis and cost effectiveness analysis). The details of the search concepts can be obtained from S1 and S2 Tables. All citations were imported into an electronic database (Endnote version X8) (23), where duplicates were removed. Studies were then uploaded into Rayyan (https://www.rayyan.ai/) for title and abstract screening. The inclusion criteria comprised of empirical studies that reported on an economic evaluation with a focus on caregiver interventions for neurodevelopmental disorders (see Table 1), in all geographical locations as summarized in Table 1. Studies that focused on physical and sensory impairments in children and studies that did not report on economic evaluation models of caregiver interventions for NDDs were excluded. Further, studies published beyond the time frame, were reviews, and in languages other than English were also excluded. The reason for exclusion of each study were noted. Two reviewers (AK and ED) independently screened titles and abstracts to assess relevance based on inclusion and exclusion criteria (Table 1). A third reviewer (EB) resolved any variations established. Abstracts included were then assessed for full-text inclusion. Full text articles eligible for inclusion were then selected for data extraction.

**Table 1:** Inclusion and exclusion criteria summary.

| Inclusion criteria | Exclusion criteria |
| --- | --- |
| 1. Studies on economic evaluation of caregiver interventions for NDDs.<br>2. Studies reporting on economic outcome measures for caregivers and/or children with DDs.<br>3. NDDs: impairments in social communication domains including autism spectrum disorder (ASD), attention deficit hyperactivity disorder (ADHD), intellectual disabilities, language communication disorders.<br>4. All countries.<br>5. Empirical studies. | 1. Studies that did not report economic evaluations of caregiver interventions for NDDs.<br>2. Studies not reporting on economic outcome measures for caregivers and/or children with DDs.<br>3. DDs: impairments: physical and sensory impairments.<br>4. Studies for interventions for caregivers of adults with disabilities<br>5. Any reviews of economic evaluations. |

### Data extraction

The data was extracted using a tailored Microsoft Excel worksheet and summarized in a narrative format: author/year, country, setting, population, type of DD, study design, intervention, comparator, and the economic evaluation methods reported guided by the Consolidated Health Economic Evaluation Reporting Standards (CHEERS) checklist (24), and the Gates Reference Case for Economic Evaluations (25). The detailed CHEERS checklist and Gates Reference Case are available in S3 Table. The extraction sheet was piloted for completeness using five sample studies. Data extraction was undertaken by AK and ED. Discrepancies in the study selection and data extraction were resolved through discussion between two reviewers (AK, ED) and a third reviewer (EB).

### Data synthesis

Economic findings were synthesised and presented as a narrative summary in conjunction with a tabular summary. Our synthesis focused on how each economic evaluation has been described in the papers and summarized the results of the studies and methodological challenges encountered in economic evaluations of caregiver interventions for children with DDs.

### Quality assessment

The quality of the included economic evaluations was rated using the Drummond 10-point checklist (26). The checklist guides on the critique of economic evaluations by considering: 1) the research question; 2) the study/intervention description; 3) the study design; 4) the identification, 5) measurement, and 6) valuation of costs and consequences; 7) application of discounting; 8) incremental analysis; 9) clear presentation of results with uncertainty and sensitivity analyses; and 10) discussion of results in the context of policy relevance and existing literature. Based on this, the studies were rated using a scale developed by Doran (27) with a potential score of 1 to each of the checklist items. The aggregate scores reflected an economic appraisal of poor quality (scores 1 to 3), average quality (scores 4 to 7) and good quality (scores 8 to 10). Authors AK and ED conducted independent quality appraisal of the included studies.

## Results

### Article selection

The literature search identified 7,811 articles. After excluding duplicate studies, 7,255 studies remained for title and abstract screening. The screening based on title and abstract resulted in 38 articles eligible for full text screening. Most studies were excluded because they included a clinical condition with no NDD, they were not primary studies reporting economic evaluations results (e.g. reviews) or focused on clinical treatment rather than caregiver interventions for children with NDDs. After full-text screening, 17 studies were included for data extraction and quality assessment. Additional details are presented in the PRISMA flow diagram (Figure 1).

**Figure 1:**
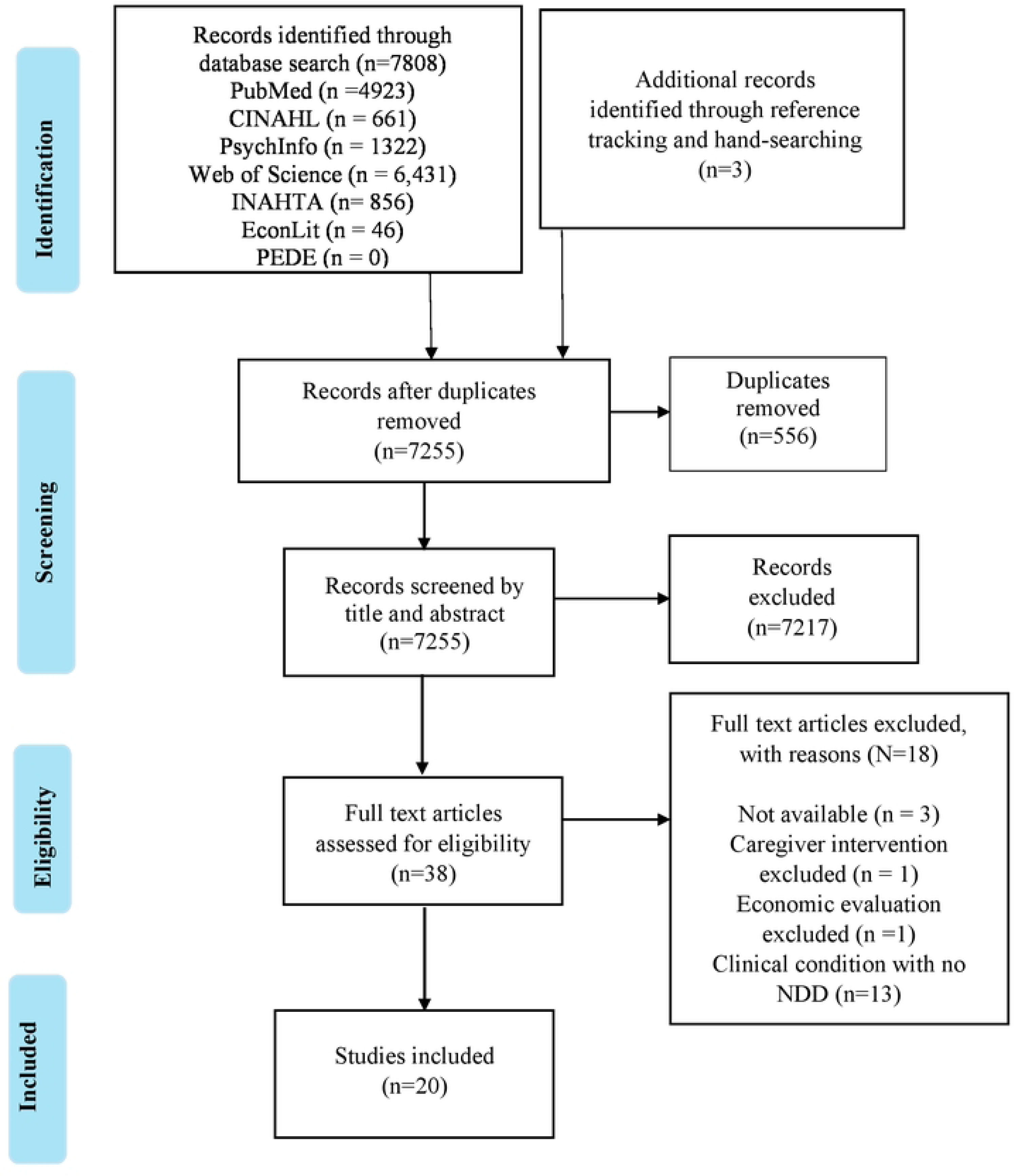
PRISMA flow chart of the study selection process.

### Quality assessment

Most studies were classified as good quality (n=16) (28–40) and four studies were classified as average (41–44). The most common reason for studies not receiving full points was due to lack of inclusion of discounting (n=12) (28, 29, 32, 34, 35, 37, 40–45) and unclear presentation and discussion of results (n=3) (31, 43, 44). Details of quality scores for each study is presented in S4 Table.

### Overview of key characteristics

All the studies included were conducted in high income countries specifically UK (n=7), Canada (n = 2), United States (n=3), Australia (n=1), Denmark (n=1), Sweden (n=3), Ireland (n=1), Japan (n=1), and multi-country (England, Ireland, Italy, Spain) (n=1). Most of the studies focused on caregiver interventions for children with autism spectrum disorder (n=7), followed by attention deficit hyperactivity disorder (ADHD) (n=6), disruptive behaviour with ADHD (n=2), behavioural problems with ADHD (n=3), intellectual disabilities (n=1) and language delay (n=1). The caregiver interventions were delivered through group format (n=9), on individual basis (n=8) or both platforms (n=1). Two of the studies did not report on the intervention delivery method. Overall, the caregiver interventions targeted children ranging from 12 months to 12 years.

Of the reviewed studies, eleven reported a cost effectiveness analysis (CEA) (28, 29, 31, 32, 35, 37, 38, 40, 41, 46, 47), two studies carried out a cost utility analysis (CUA) (33, 44), one study performed a cost consequence analysis (48) and one study conducted a cost benefit analysis (CBA) (36). However, two studies reported as CEAs (31, 38) were CUAs based on the measure of outcome as quality adjusted life years (QALYs) and one study conducted an additional CUA (46). In addition to these, two studies conducted combined analyses as a cost analysis alongside a CEA (30), and a cost consequence analysis (CCA) alongside a CEA (39). For partial economic evaluations, three studies conducted a costing analysis (34, 42, 43). Fourteen were economic evaluations conducted alongside trials (28, 29, 31, 32, 34, 38–44, 46, 47), whereas five studies were model based (30, 33, 36, 37, 48). One study did not state the economic evaluation approach used (35).The time horizon for the majority of studies was relatively short, that is: 13 weeks for one study (41); six to ten months for three studies (29, 34, 42); and, one year for two studies (32, 46). Eight studies extrapolated to a long-term period ranging from two years to 65 years (30, 33, 35–39, 48). Six studies (28, 31, 40, 43, 44, 47) did not explicitly state the time horizon of the evaluations. In Table 2 we provide a detailed overview of the main characteristics of the included studies.

**Table 2:** Overview of the key characteristics of the included economic evaluation studies.

| Author & year | Developmental disability | Country | Study population (age group) | Study design (Model type) | Type of economic evaluation | Follow-up period (trials) or time horizon (models) |
| --- | --- | --- | --- | --- | --- | --- |
| Scavenius et al (28), 2020 | Attention-deficit hyperactivity disorder (ADHD) | Denmark | 3 to 9 years | Randomized controlled trial | Cost effectiveness analysis | Not reported |
| Sayal et al (29), 2016 | Attention-deficit hyperactivity disorder (ADHD) | United Kingdom | 4 to 8 years | Three arm randomized cluster trial | Cost effectiveness analysis | 9 months (inferred) |
| Segal et al (30), 2023 | Autism Spectrum Disorder | Australia | 12 months | Decision analytic model (Decision tree) | Cost analysis and Cost-effectiveness analysis | 12 years |
| Sonuga-Barke et al<br>(44), 2017 | Attention-deficit<br>hyperactivity<br>disorder (ADHD) | United<br>Kingdom | 2 years 9 months to<br>4 years 6 months | Multi-centre three-arm<br>parallel randomised<br>controlled | Cost analysis (initially<br>planned to do a cost<br>utility analysis) | Not reported |
| Charman et al<br>(31), 2021 | Autism Spectrum<br>Disorder | United<br>Kingdom | 4 to 8.11 years | Randomized controlled<br>trial | Cost effectiveness<br>analysis | Not reported |
| Byford et al (32),<br>2015 | Autism Spectrum<br>Disorder | United<br>Kingdom | 2 to 4 years 11<br>months | Multi centre randomised<br>controlled trial | Cost effectiveness<br>analysis | 13 months |
| Nystrand et al (33),<br>2019 | Behaviour problems<br>(conduct disorder<br>and ADHD) | Sweden | 5 to 12 years | Decision analytic model<br>(Markov) | Cost utility analysis | 18 years |
| Tsiplova et al (34),<br>2022 | Autism Spectrum<br>Disorder | Canada | 15 to 36 months | Randomized comparative<br>effectiveness trial | Cost analysis | 24 weeks |
| Kuklinski et al<br>(35), 2023 | Disruptive<br>behaviour (ADHD) | United States | Not reported | Economic evaluation | Cost effectiveness<br>analysis | 5 years |
|  | and conduct disorder) |  |  |  |  |  |
| Nystrand et al (36),<br>2019 | Behaviour problems (conduct disorder and ADHD) | Sweden | 5 to 12 years | Decision analytic model (Markov) | Cost benefit analysis | 2 years |
| Tran et al (41),<br>2018 | Attention-deficit hyperactivity disorder (ADHD) | United States | 7 to 11 years | Randomized controlled trial | Cost effectiveness analysis | 13 weeks |
| Page et al (42),<br>2016 | Attention-deficit hyperactivity disorder (ADHD) | United States | 5 to 12 years | Randomized controlled trial | Cost analysis | 10 months |
| Penner et al (37),<br>2015 | Autism Spectrum Disorder | Canada | 15 to 36 months | Decision analytic model (Decision tree) | Cost effectiveness analysis | 65 years |
| Nystrand et al (38),<br>2020 | Disruptive behaviour | Sweden | 8 to 12 years | Randomized controlled trial | Cost effectiveness analysis | 2 years |
| Ferguson et al (43),<br>2022 | Autism Spectrum<br>Disorder | Ireland | 2 to 6 years | Experimental research<br>design (trial) | Cost analysis | Not reported |
| O'Farrelly et al<br>(39), 2021 | Behaviour problems | United<br>Kingdom | 12 to 36 months | Randomised controlled<br>trial | Cost effectiveness<br>analysis; cost<br>consequence analysis | Short term analysis<br>24 months; Long<br>term analysis<br>beyond trial period |
| Gibbard et al (40),<br>2004 | Language delay | United<br>Kingdom | 22 to 36 months | Randomised controlled<br>trial | Cost effectiveness<br>analysis | Not reported |
| Royston et al (46) | Intellectual<br>disabilities | United<br>Kingdom | 30 to 59 months | Randomised controlled<br>trial | Cost effectiveness<br>analysis (and cost<br>utility analysis) | 12 months |
| Tinelli et al (48) | Autism Spectrum<br>Disorder | Multi-country<br>(England,<br>Ireland, Italy,<br>Spain) | 2 years to 4 years 11<br>months | Economic evaluation | Cost consequence<br>analysis | 6 years |
| Shimabukuro et al<br>(47) | Attention-deficit<br>hyperactivity<br>disorder (ADHD) | Japan | 6 to 12 years | Randomised controlled<br>trial | Cost effectiveness<br>analysis | Not reported |

### Economic evaluation methods

#### Cost estimates

We summarized the cost estimates associated with caregiver interventions implementation and the associated healthcare costs for children with DDs. Nine studies conducted evaluations from one perspective only: four studies adopted a health provider’s perspective (28, 33, 35, 43); two studies adopted a government (insurer) payer perspective (30, 39); and three studies adopted a societal perspective (38, 40, 41). Nine studies reported results from more than one perspective (31, 32, 34, 37, 40, 44, 46–48). The remaining two studies did not explicitly state the perspective adopted (36, 42).

The health provider costs mainly reported on the intervention implementation costs, for instance training, personnel, overheads, treatment costs amongst others. Among these, only three studies (28, 35, 46) reported on both start-up and implementation costs of the caregiver interventions. Notably, Scavenius et al (2020) reported only financial costs and excluded opportunity costs such as income loss or other sector benefits, as they were not tangible costs of the intervention implementation (28). Cost inclusion or exclusion may impact the study results by increasing the costs of the intervention, but at the same time may also reduce the incremental costs of the intervention. Four studies (33, 36, 44, 46) captured the costs of other sectors i.e. education, social and crime services. Furthermore, Nystrand et al (2019; 2020) included cost offsets as the treatment costs avoided with the reduction of the disease prevalence incurred by the health sector as a cost input (33, 36). For societal costs, four studies (37, 38, 46, 47) mainly included either direct costs (health service costs, medication, childcare, travel) or indirect costs (productivity loss), whereas eight studies (29, 31, 32, 34, 40, 41, 44, 48) comprehensively captured both direct and indirect costs. Most studies explicitly stated the currency and reported year used except six studies (29, 31, 32, 42–44). Discounting was not necessary in the studies which collected costs over a one-year time horizon or less. However, where necessary due to a longer time horizon adopted by twelve of the included studies, a discount rate ranging from 2% to 6% per annum was applied with the exception of two studies (35, 37). Therefore, the cost-effectiveness results reported may be inaccurate for both studies.

### Health outcome measures

We report key effectiveness outcomes of caregiver interventions and comparators in the eligible studies in Table 4. Except for the cost analysis, the most frequently reported health outcomes for cost utility analysis included four studies reporting on QALYs (29, 31, 38, 46), and one study studies on disability adjusted life years (DALYs) (33). Of the studies that reported QALYs, the generic health related quality of life (HRQoL) measures used were Child Health Utility 9D (CHU-9D) and EuroQoL 5 dimensions Youth (EQ5D-Y). Health outcomes reported in 9 studies applying cost effectiveness analysis were mostly per case reduction and per outcome score improvement amongst others (28, 30, 32, 35, 39–41, 46, 47). O’Farrelly et al (2021) carried out a short term CEA and a long term CUA, but with the small differences in the QALYs and no difference in cost between the trial groups, only the short term CEA was reported with per outcome score improvement as the health outcome (39). Further, Penner et al (2015) reported dependency-free life years (DFLYs) as the outcome measure (37), while Nystrand et al (2020) reported the net present value for the CBA (36). Additional details included in each of the reviewed studies are listed in Table 3.

**Table 3:**
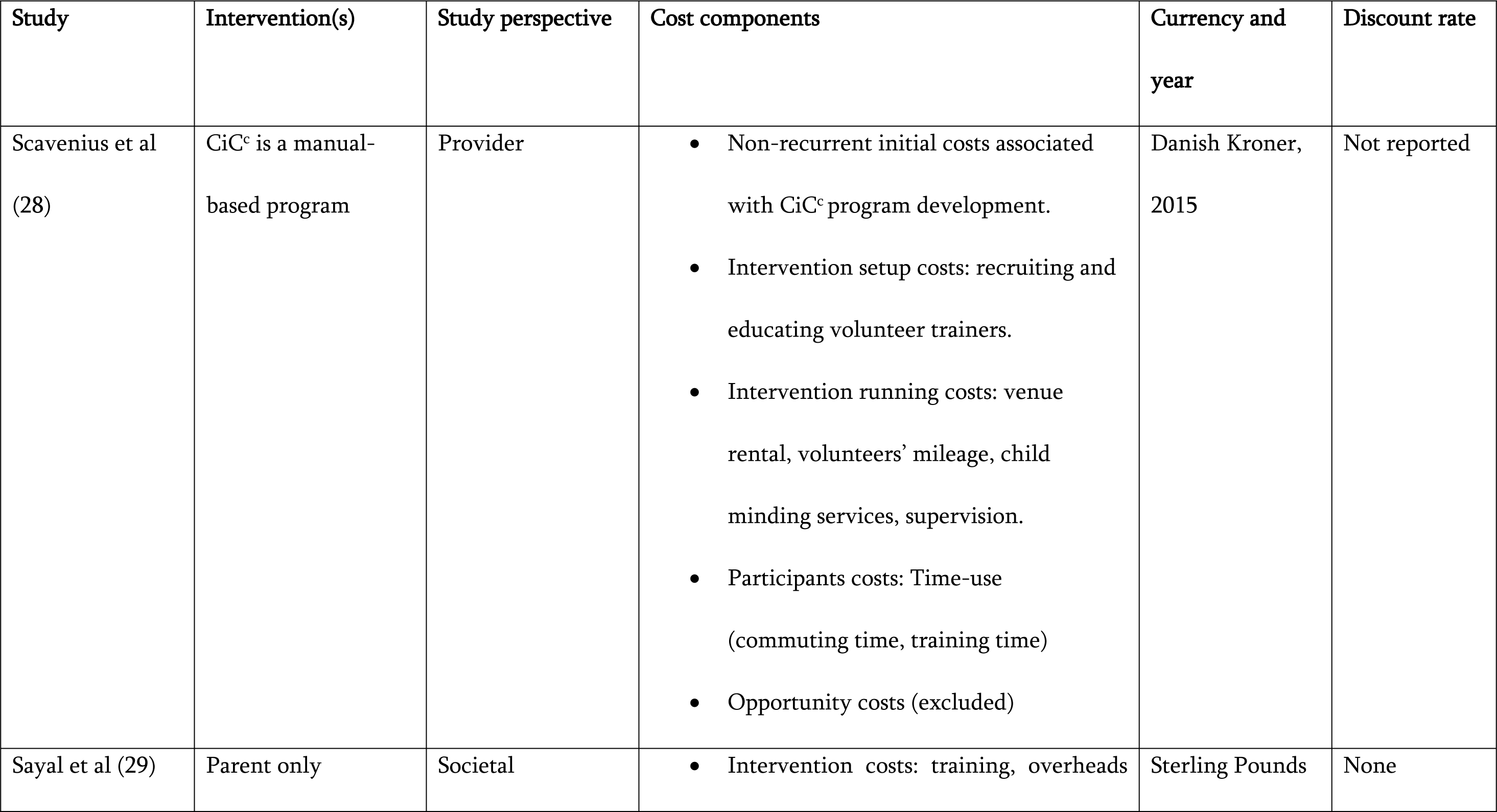
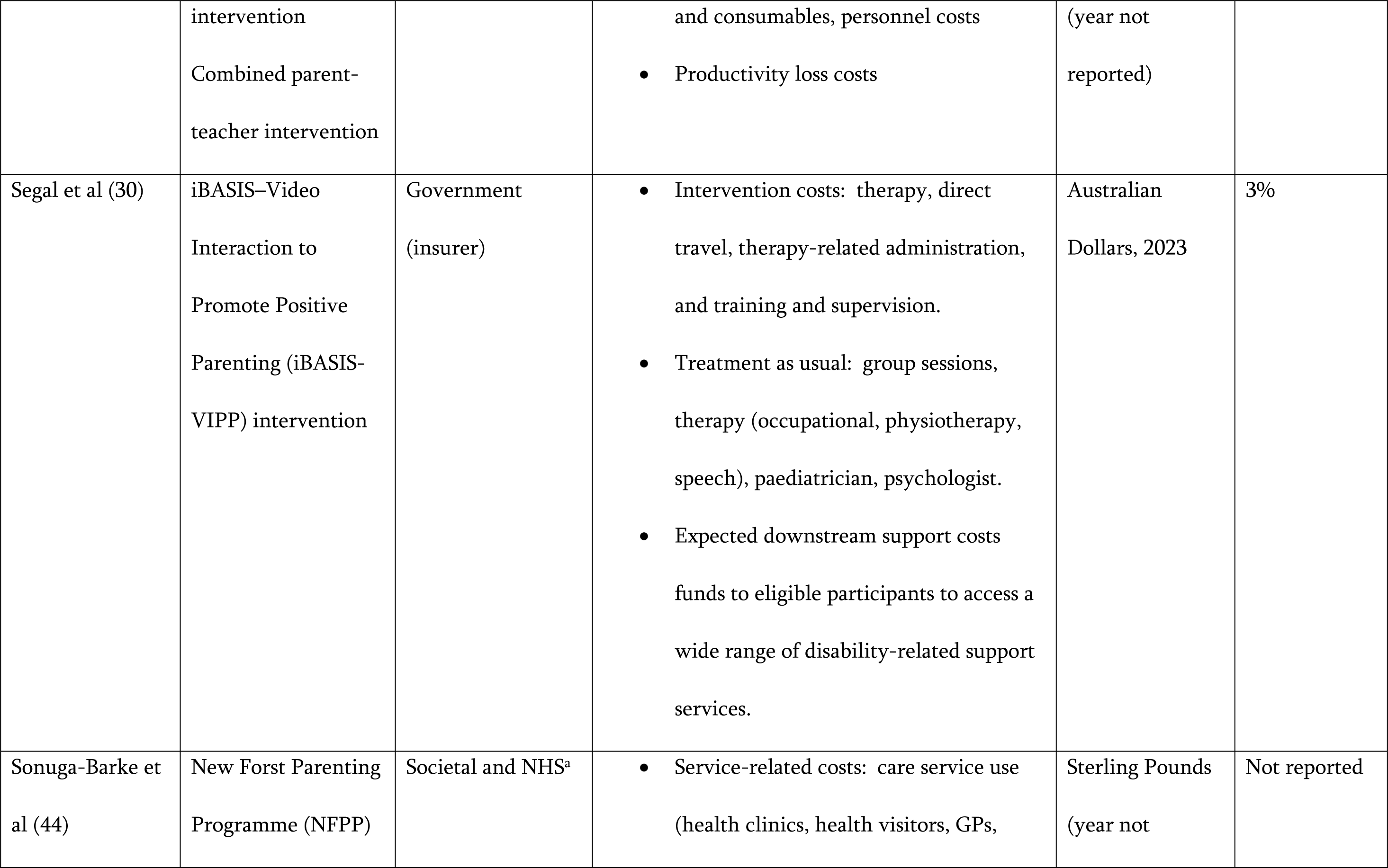
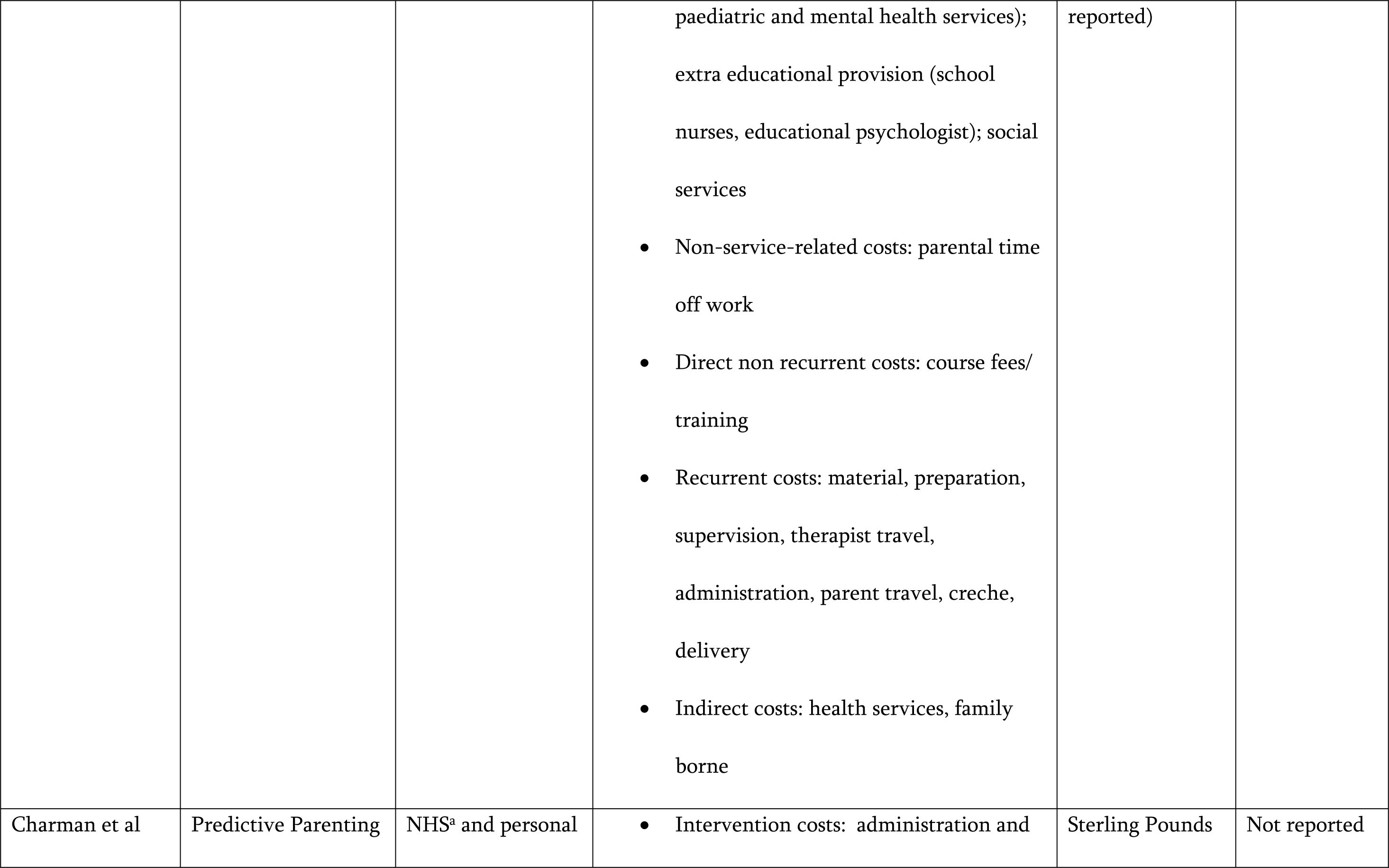
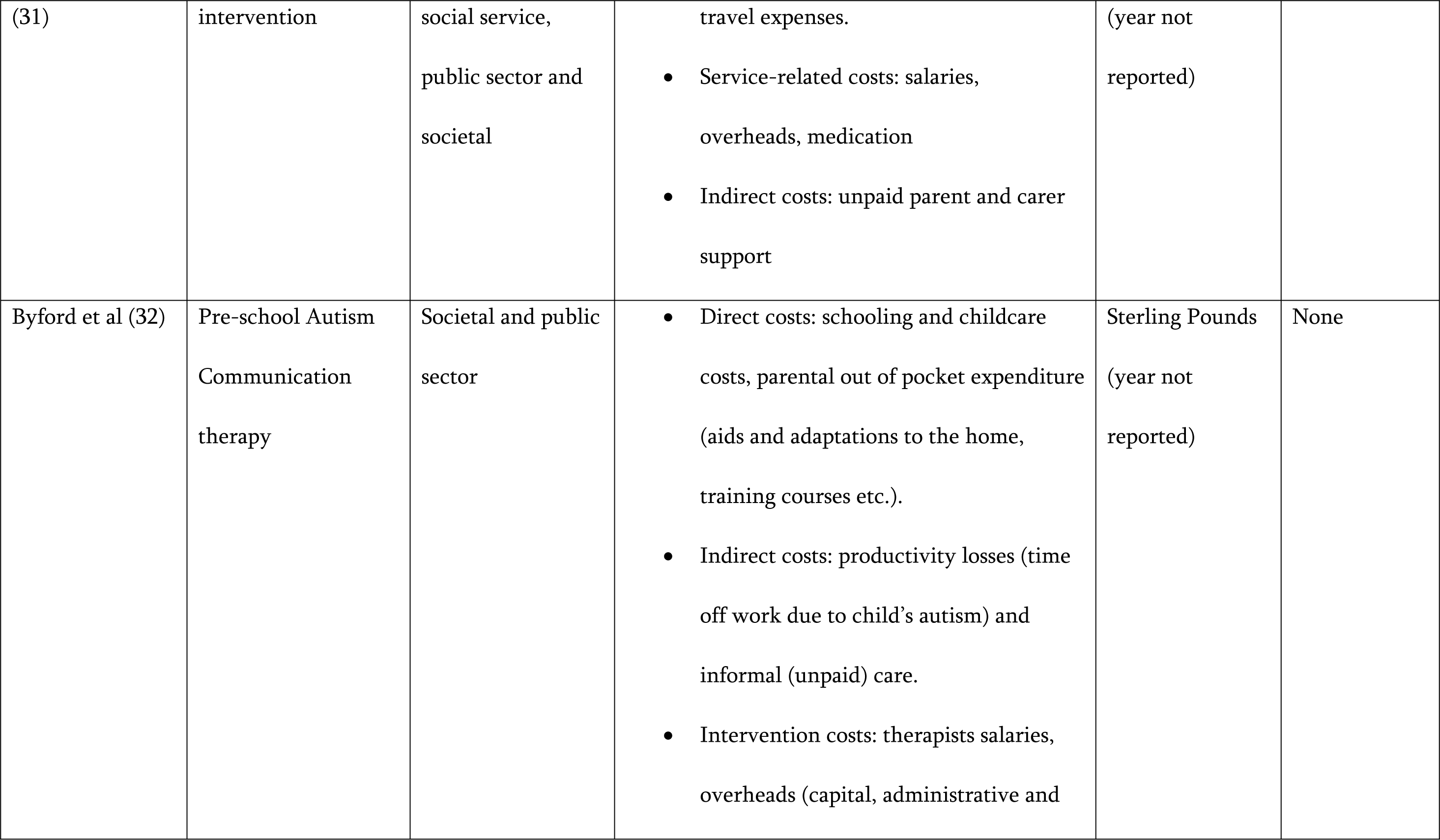
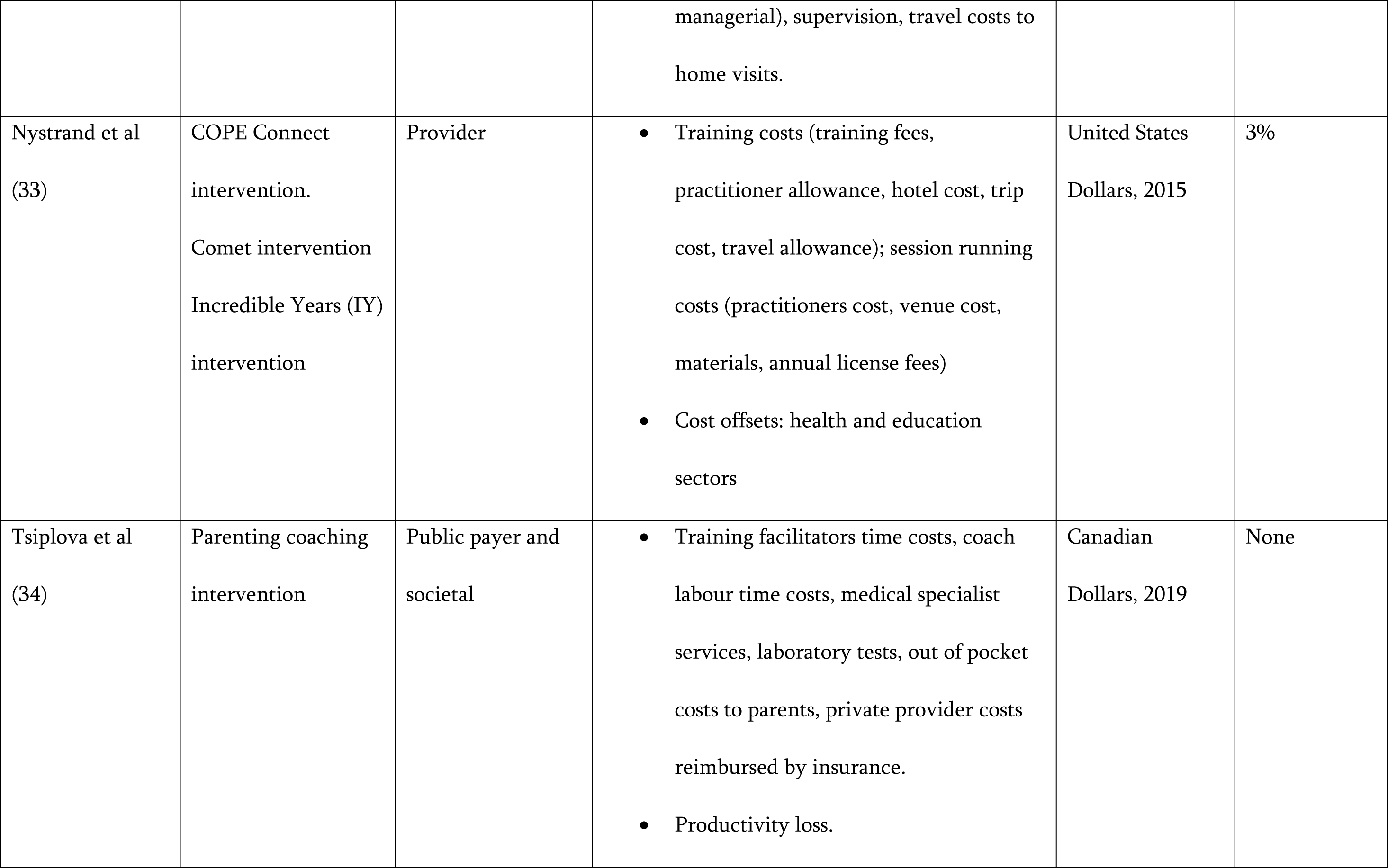
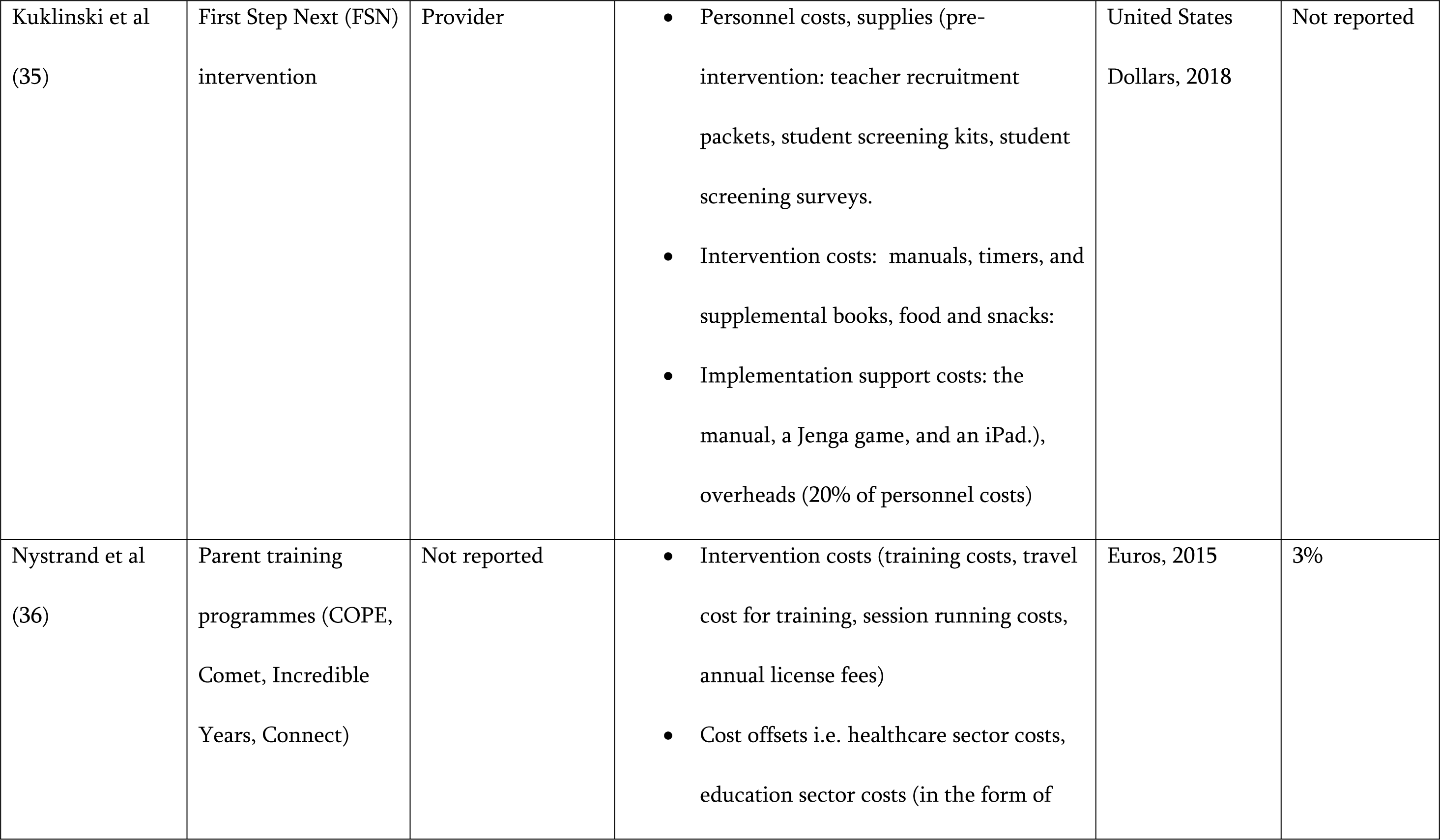
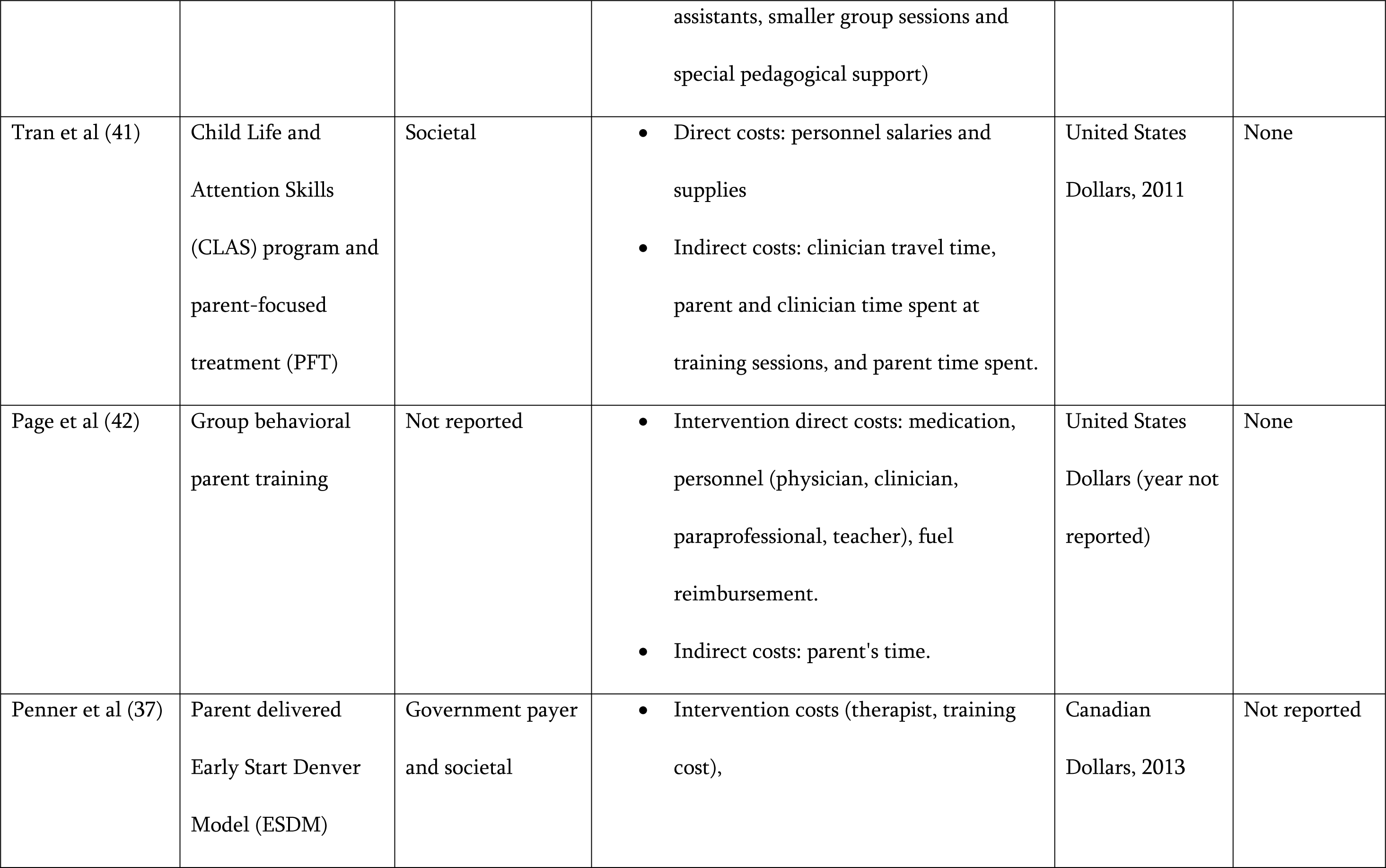
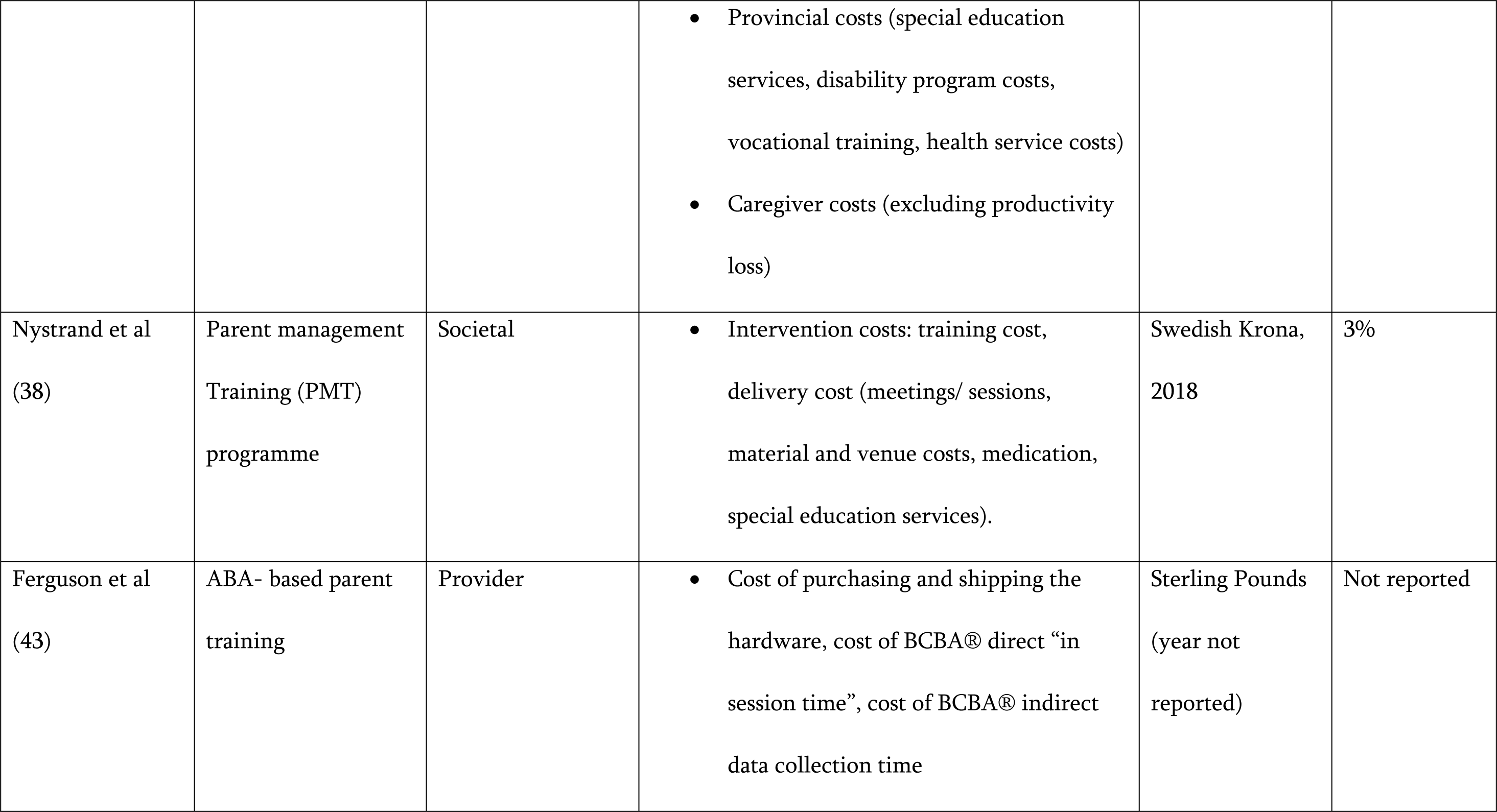
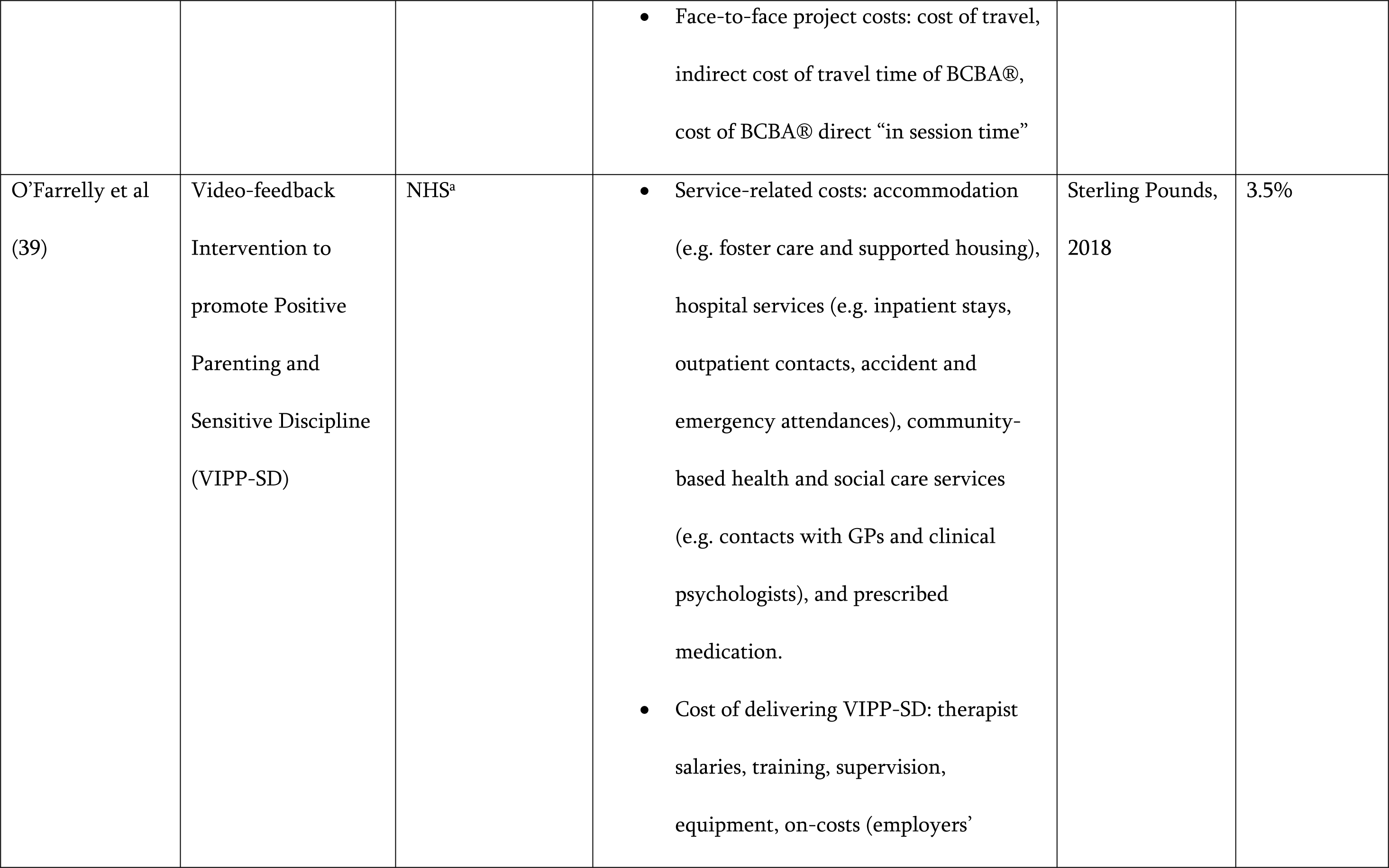
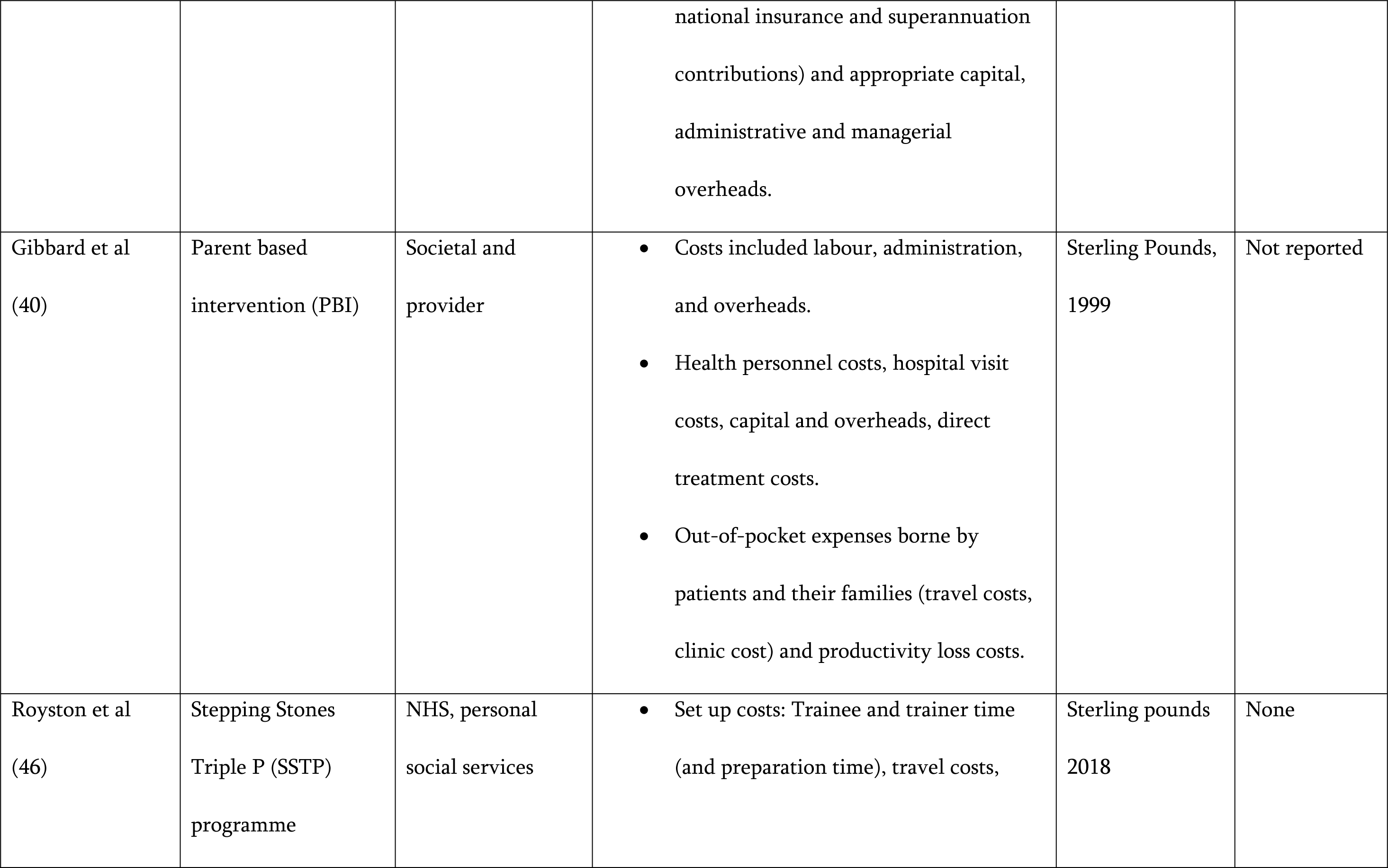
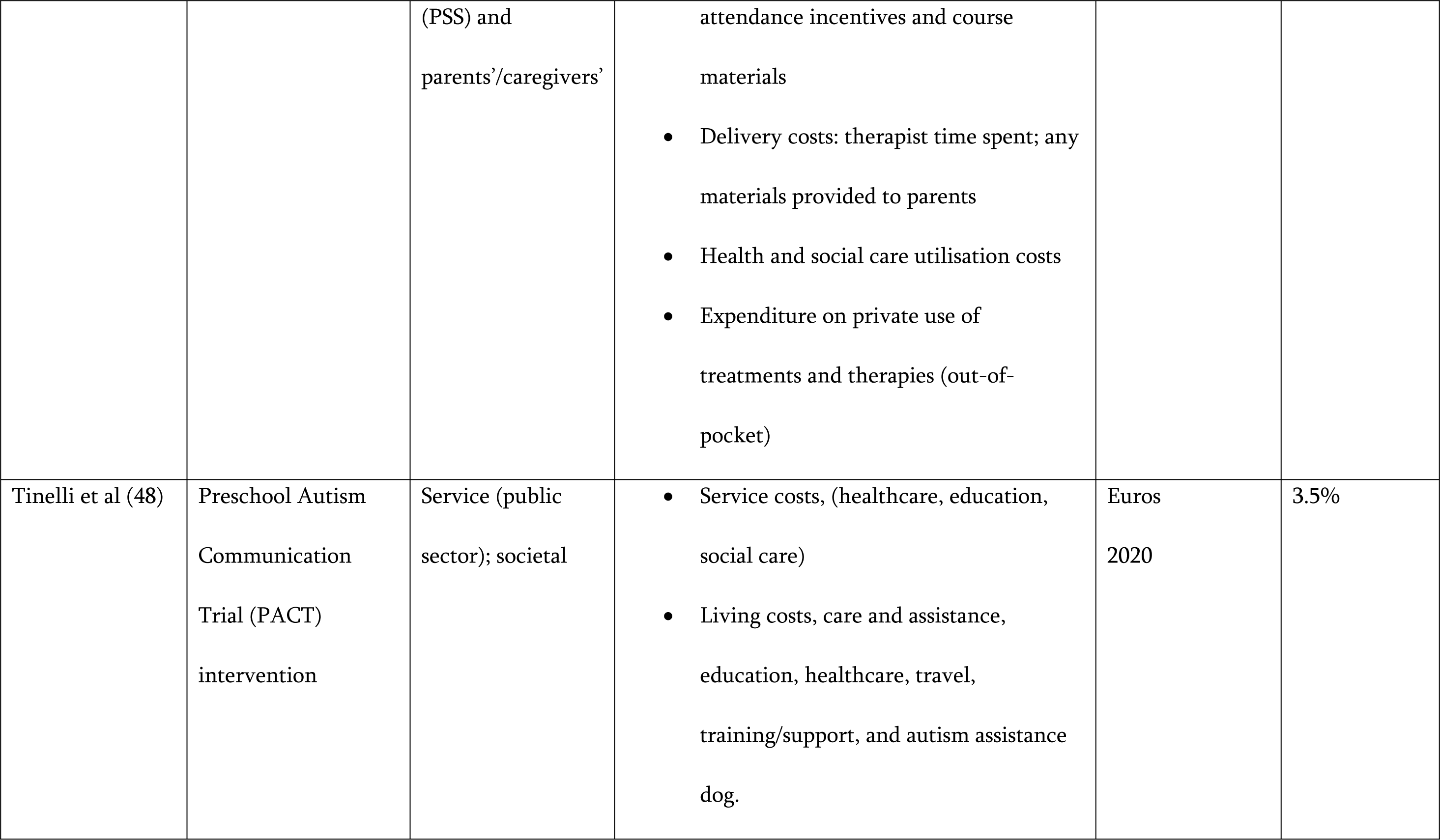
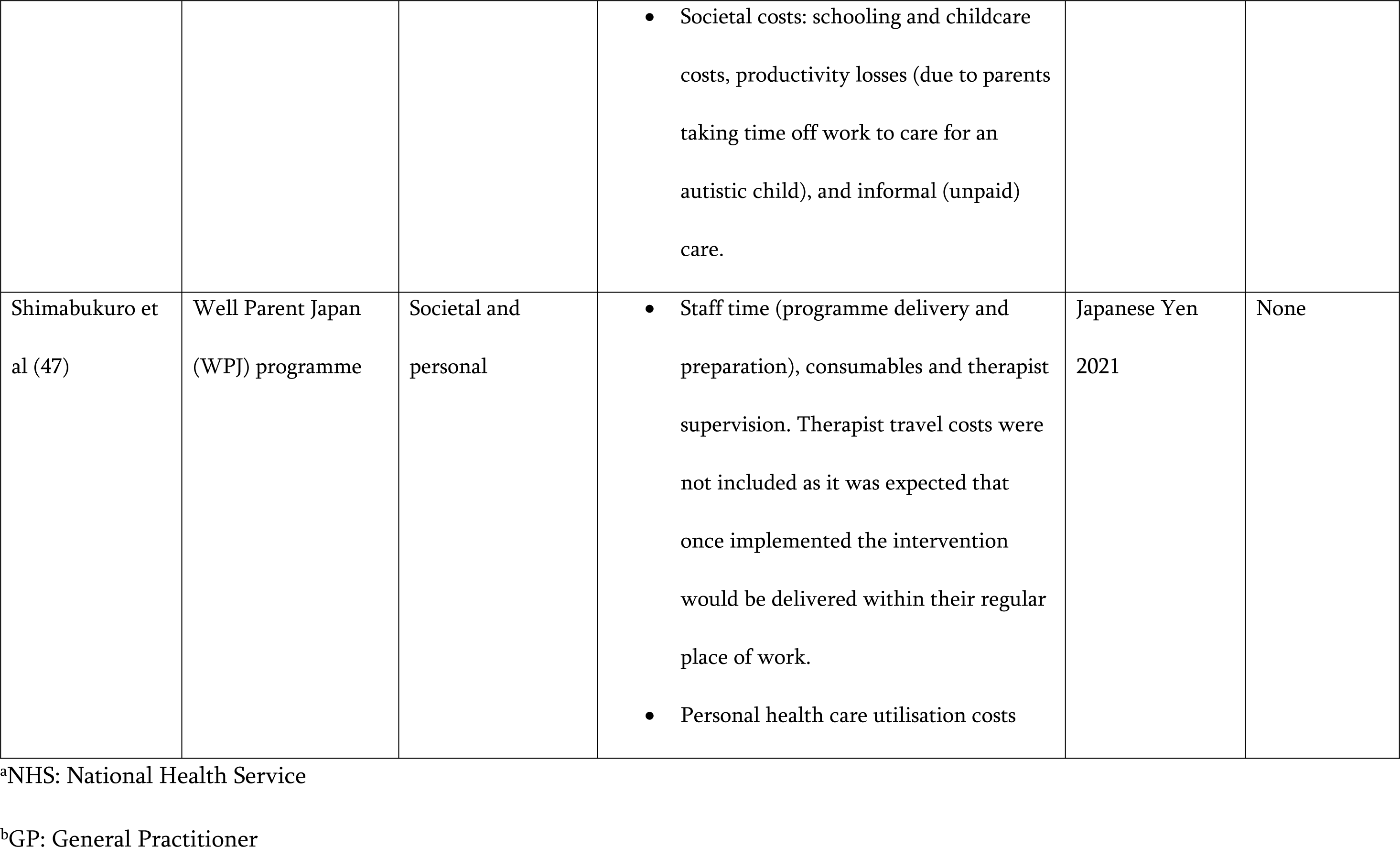
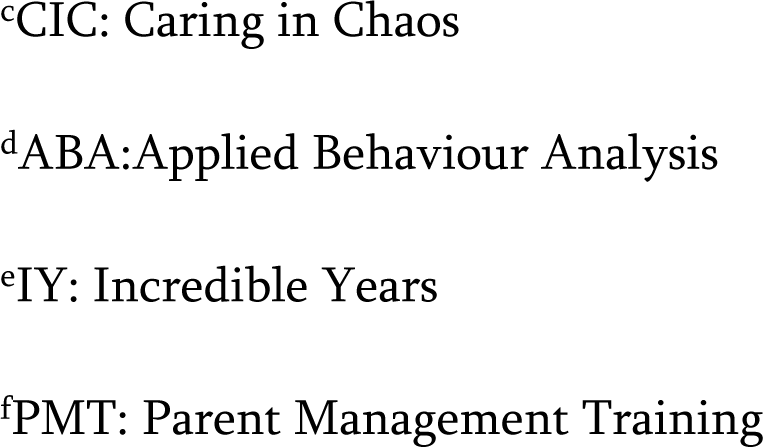
Cost inputs associated with the caregiver interventions included in this review.

| Study | Intervention(s) | Study perspective | Cost components | Currency and year | Discount rate |
| --- | --- | --- | --- | --- | --- |
| Scavenius et al (28) | CiC<sup>c</sup> is a manual-based program | Provider | • Non-recurrent initial costs associated with CiC<sup>c</sup> program development.<br>• Intervention setup costs: recruiting and educating volunteer trainers.<br>• Intervention running costs: venue rental, volunteers' mileage, child minding services, supervision.<br>• Participants costs: Time-use (commuting time, training time)<br>• Opportunity costs (excluded) | Danish Kroner, 2015 | Not reported |
| Sayal et al (29) | Parent only | Societal | • Intervention costs: training, overheads | Sterling Pounds | None |
|  | intervention<br>Combined parent-teacher intervention |  | and consumables, personnel costs<br>• Productivity loss costs | (year not<br>reported) |  |
| Segal et al (30) | iBASIS–Video<br>Interaction to<br>Promote Positive<br>Parenting (iBASIS-VIPP) intervention | Government<br>(insurer) | • Intervention costs: therapy, direct travel, therapy-related administration, and training and supervision.<br>• Treatment as usual: group sessions, therapy (occupational, physiotherapy, speech), paediatrician, psychologist.<br>• Expected downstream support costs funds to eligible participants to access a wide range of disability-related support services. | Australian<br>Dollars, 2023 | 3% |
| Sonuga-Barke et al (44) | New Forst Parenting<br>Programme (NFPP) | Societal and NHS<sup>a</sup> | • Service-related costs: care service use (health clinics, health visitors, GPs, | Sterling Pounds<br>(year not | Not reported |
|  |  |  | paediatric and mental health services);<br>extra educational provision (school<br>nurses, educational psychologist); social<br>services<br>• Non-service-related costs: parental time<br>off work<br>• Direct non recurrent costs: course fees/<br>training<br>• Recurrent costs: material, preparation,<br>supervision, therapist travel,<br>administration, parent travel, creche,<br>delivery<br>• Indirect costs: health services, family<br>borne | reported) |  |
| Charman et al | Predictive Parenting | NHS<sup>a</sup> and personal | • Intervention costs: administration and | Sterling Pounds | Not reported |
| (31) | intervention | social service,<br>public sector and<br>societal | travel expenses.<br>• Service-related costs: salaries, overheads, medication<br>• Indirect costs: unpaid parent and carer support | (year not reported) |  |
| Byford et al (32) | Pre-school Autism<br>Communication<br>therapy | Societal and public<br>sector | • Direct costs: schooling and childcare costs, parental out of pocket expenditure (aids and adaptations to the home, training courses etc.).<br>• Indirect costs: productivity losses (time off work due to child's autism) and informal (unpaid) care.<br>• Intervention costs: therapists salaries, overheads (capital, administrative and<br>managerial), supervision, travel costs to home visits. | Sterling Pounds<br>(year not reported) | None |
| Nystrand et al<br>(33) | COPE Connect intervention.<br>Comet intervention<br>Incredible Years (IY) intervention | Provider | • Training costs (training fees, practitioner allowance, hotel cost, trip cost, travel allowance); session running costs (practitioners cost, venue cost, materials, annual license fees)<br>• Cost offsets: health and education sectors | United States Dollars, 2015 | 3% |
| Tsiplova et al<br>(34) | Parenting coaching intervention | Public payer and societal | • Training facilitators time costs, coach labour time costs, medical specialist services, laboratory tests, out of pocket costs to parents, private provider costs reimbursed by insurance.<br>• Productivity loss. | Canadian Dollars, 2019 | None |
| Kuklinski et al<br>(35) | First Step Next (FSN) intervention | Provider | • Personnel costs, supplies (pre-intervention: teacher recruitment packets, student screening kits, student screening surveys.<br>• Intervention costs: manuals, timers, and supplemental books, food and snacks:<br>• Implementation support costs: the manual, a Jenga game, and an iPad.), overheads (20% of personnel costs) | United States<br>Dollars, 2018 | Not reported |
| Nystrand et al<br>(36) | Parent training programmes (COPE, Comet, Incredible Years, Connect) | Not reported | • Intervention costs (training costs, travel cost for training, session running costs, annual license fees)<br>• Cost offsets i.e. healthcare sector costs, education sector costs (in the form of<br>assistants, smaller group sessions and special pedagogical support) | Euros, 2015 | 3% |
| Tran et al (41) | Child Life and Attention Skills (CLAS) program and parent-focused treatment (PFT) | Societal | • Direct costs: personnel salaries and supplies<br>• Indirect costs: clinician travel time, parent and clinician time spent at training sessions, and parent time spent. | United States Dollars, 2011 | None |
| Page et al (42) | Group behavioral parent training | Not reported | • Intervention direct costs: medication, personnel (physician, clinician, paraprofessional, teacher), fuel reimbursement.<br>• Indirect costs: parent's time. | United States Dollars (year not reported) | None |
| Penner et al (37) | Parent delivered Early Start Denver Model (ESDM) | Government payer and societal | • Intervention costs (therapist, training cost), | Canadian Dollars, 2013 | Not reported |
|  |  |  | Provincial costs (special education services, disability program costs, vocational training, health service costs)<br>Caregiver costs (excluding productivity loss) |  |  |
| Nystrand et al<br>(38) | Parent management<br>Training (PMT)<br>programme | Societal | Intervention costs: training cost, delivery cost (meetings/ sessions, material and venue costs, medication, special education services). | Swedish Krona,<br>2018 | 3% |
| Ferguson et al<br>(43) | ABA- based parent<br>training | Provider | Cost of purchasing and shipping the hardware, cost of BCBA® direct “in session time”, cost of BCBA® indirect data collection time | Sterling Pounds<br>(year not<br>reported) | Not reported |
|  |  |  | • Face-to-face project costs: cost of travel, indirect cost of travel time of BCBA®, cost of BCBA® direct “in session time” |  |  |
| O’Farrelly et al<br>(39) | Video-feedback<br>Intervention to<br>promote Positive<br>Parenting and<br>Sensitive Discipline<br>(VIPP-SD) | NHS<sup>a</sup> | • Service-related costs: accommodation (e.g. foster care and supported housing), hospital services (e.g. inpatient stays, outpatient contacts, accident and emergency attendances), community-based health and social care services (e.g. contacts with GPs and clinical psychologists), and prescribed medication.<br>• Cost of delivering VIPP-SD: therapist salaries, training, supervision, equipment, on-costs (employers’ | Sterling Pounds,<br>2018 | 3.5% |
|  |  |  | national insurance and superannuation contributions) and appropriate capital, administrative and managerial overheads. |  |  |
| Gibbard et al<br>(40) | Parent based intervention (PBI) | Societal and provider | Costs included labour, administration, and overheads.<br>Health personnel costs, hospital visit costs, capital and overheads, direct treatment costs.<br>Out-of-pocket expenses borne by patients and their families (travel costs, clinic cost) and productivity loss costs. | Sterling Pounds, 1999 | Not reported |
| Royston et al<br>(46) | Stepping Stones Triple P (SSTP) programme | NHS, personal social services | Set up costs: Trainee and trainer time (and preparation time), travel costs, | Sterling pounds 2018 | None |
|  |  | (PSS) and<br>parents'/caregivers' | attendance incentives and course materials<br>• Delivery costs: therapist time spent; any materials provided to parents<br>• Health and social care utilisation costs<br>• Expenditure on private use of treatments and therapies (out-of-pocket) |  |  |
| Tinelli et al (48) | Preschool Autism<br>Communication<br>Trial (PACT)<br>intervention | Service (public<br>sector); societal | • Service costs, (healthcare, education, social care)<br>• Living costs, care and assistance, education, healthcare, travel, training/support, and autism assistance dog. | Euros<br>2020 | 3.5% |
|  |  |  | Societal costs: schooling and childcare costs, productivity losses (due to parents taking time off work to care for an autistic child), and informal (unpaid) care. |  |  |
| Shimabukuro et al (47) | Well Parent Japan (WPJ) programme | Societal and personal | Staff time (programme delivery and preparation), consumables and therapist supervision. Therapist travel costs were not included as it was expected that once implemented the intervention would be delivered within their regular place of work.<br>Personal health care utilisation costs | Japanese Yen 2021 | None |

<sup>a</sup>NHS: National Health Service
<sup>b</sup>GP: General Practitioner
<sup>c</sup>CIC: Caring in Chaos
<sup>d</sup>ABA: Applied Behaviour Analysis
<sup>e</sup>IY: Incredible Years
<sup>f</sup>PMT: Parent Management Training

**Table 4:** Clinical and economic outcomes of the economic evaluation studies included in this review.

| Study | Intervention(s) | Comparator | Clinical outcome | Economic outcome | Sensitivity analysis |
| --- | --- | --- | --- | --- | --- |
| **Cost-effectiveness analysis** |  |  |  |  |  |
| Scavenius et al (28) | CiC<sup>a</sup> is a manual-based program | Waitlist control group | Parenting competence measure by Parenting Sense Competence Scale (PSOC)<br>Child functioning measured by Home Situations Questionnaire (HSQ) | per Cohen's d effect sizes (ES) improvement (in the PSOC and HSQ) | Not reported |
| Segal et al (30) | iBASIS–Video Interaction to Promote Positive Parenting (iBASIS-VIPP)<br>intervention | Treatment as usual | Incidence of ASD diagnosis | per lower incident case of diagnosed ASD | Univariate analyses and probabilistic analyses |
| Byford et al<br>(32) | Pre-school Autism<br>Communication therapy | Treatment as usual | (ADOS-G) Autism<br>Diagnostic Observation<br>Schedule-Generic score | per ADOS-G score<br>improvement | Not reported |
| Kuklinski et al<br>(35) | First Step Next (FSN)<br>intervention | Home based<br>intervention (hB) | Clinical outcomes (ADHD<sup>b</sup><br>vs CD<sup>d</sup> vs ADHD<sup>b</sup> plus CD<sup>d</sup>) | Per case reduction | Probabilistic sensitivity<br>analysis |
| Tran et al (41) | Child Life and Attention<br>Skills (CLAS) program<br>and parent-focused<br>treatment (PFT) | Treatment as usual | Not reported | Per resolved ADHD<sup>b</sup>-I<br>case | One way and multiway<br>sensitivity analysis |
| O'Farrelly et<br>al (39) | Video-feedback<br>Intervention to promote<br>Positive Parenting and | Treatment as usual | Preschool Parental Account<br>of Children's Symptoms<br>(PPACS) score | Per point improvement<br>in PPACS score | Probabilistic sensitivity<br>analysis and scenario<br>analysis |
|  | Sensitive Discipline<br>(VIPP-SD) |  |  | Quality adjusted life<br>years (QALYs) (not<br>reported) |  |
| Gibbard et al<br>(40) | Parent based<br>intervention (PBI) | Treatment as usual | Six measures on the child's<br>linguistic complexity from a<br>single word level to that of<br>using three-to-four-word<br>utterances | Per change in measure<br>score | Univariate sensitivity<br>analysis |
| Royston et al<br>(46) | Stepping Stones<br>Triple P (SSTP)<br>programme | Treatment as usual | Child Behaviour Check List<br>(CBCL) score | per CBCL score<br>improvement | Univariate sensitivity<br>analysis |
| Shimabukuro<br>et al (47) | Well Parent Japan (WPJ)<br>programme | Treatment as usual | Parent-domain parenting<br>stress score | per reduction in<br>parent-domain<br>parenting stress score | Not reported |
| **Cost utility analysis** |  |  |  |  |  |
| Sayal et al<br>(29) | Parent only intervention<br>Combined parent-teacher intervention | Waitlist control<br>group | • Parent- rated ADHD index<br>• Scores on the other parent-rated sub-scales and all the teacher-rated sub-scales of the Conners' Rating Scale – Revised | Quality adjusted life<br>years (QALYs) | Not reported |
| Charman et al<br>(31) | Predictive Parenting<br>intervention | Psychoeducation | • Behaviours That Challenge (BTC) per minute<br>• Autism Parenting Stress Index, Parent Efficacy scale | Quality adjusted life<br>years (QALYs) | Univariate analysis |
| Nystrand et al | COPE Connect | Waitlist control | Swanson, Nolan and | Disability adjusted life | Probabilistic sensitivity |
| (33) | intervention.<br>Comet intervention<br>Incredible Years (IY)<br>intervention | group | Pelham Scale (SNAP-IV)<br>score<br>Eyberg Child Behaviour<br>Inventory (ECBI) score | years (DALYs) | analysis and univariate<br>sensitivity analyses |
| Penner et al<br>(37) | Parent delivered Early<br>Start Denver Model<br>(ESDM) | Status Quo | Intelligence Quotient (IQ) | Dependency-free life<br>years (DFLYs) | Multiple one-way<br>sensitivity analysis |
| Nystrand et al<br>(38) | Parent management<br>Training (PMT)<br>programme | Coping Power<br>Programme and<br>parent management<br>Training (PMT)<br>programme | Parent-rated Disruptive<br>Behaviour Disorder rating<br>scale (DBD-ODD) | Quality adjusted life<br>years (QALYs) | Scenario analysis |
| Royston et al<br>(46) | Stepping Stones<br>Triple P (SSTP)<br>programme | Treatment as usual | Child Behaviour Check List<br>(CBCL) score | Quality adjusted life<br>years (QALYs) | Univariate analysis |

| Cost benefit analysis |  |  |  |  |  |
| --- | --- | --- | --- | --- | --- |
| Nystrand et al (36) | Parent training programmes (COPE, Comet, Incredible Years, Connect) | Waitlist control group | Swanson, Nolan and Pelham scale (SNAP-IV) scores | Net present value | Probabilistic sensitivity analysis |
| Other economic evaluations |  |  |  |  |  |
| Sonuga-Barke et al (44) | New Forst Parenting Programme (NFPP) | Incredible Years (IY) | Parent ratings of child's ADHD symptoms<br>Reduction of parent-rated ADHD symptoms | N/A | Not reported |
| Tsiplova et al (34) | Parenting coaching intervention | Enhanced community treatment | Not reported | N/A | Univariate analysis |
| Page et al (42) | Group behavioral parent training | Medication for ADHD<sup>b</sup> | Not reported | N/A | Univariate analyses |
| Ferguson et al<br>(43) | ABA<sup>c</sup>- based parent<br>training | Face to face model<br>training | Child affect measure | N/A | Not reported |
| Tinelli et al<br>(48) | Preschool Autism<br>Communication Trial<br>(PACT) intervention | Treatment as usual | Autism Diagnostic<br>Observation Schedule-Generic (ADOS-G) score | N/A | Deterministic<br>sensitivity analyses |

<sup>a</sup>CIC: Caring in Chaos
<sup>b</sup>ADHD: Attentive Deficit Hyperactivity Disorder
<sup>c</sup>ABA:Applied Behaviour Analysis
<sup>d</sup>CD; Conduct disorder
<sup>e</sup>PMT: Parent Management Training

### Sensitivity analysis

At least one form of sensitivity analysis was carried out in 14 of the eligible studies as shown in Table 4. Scenario analysis was carried out in two studies (38, 39), univariate sensitivity analysis was carried out in eight studies (30, 31, 33, 34, 40–42, 46), two studies (37, 41) carried out a multivariate sensitivity analysis, one study carried out a deterministic sensitivity analysis (48) and probabilistic sensitivity analysis was carried out in five studies (30, 33, 35, 36, 39). Six studies did not conduct sensitivity analysis to explore uncertainties around their results (28, 29, 32, 43, 44, 47).

### Statistical analysis

Of the trial based economic evaluations, twelve of these studies further applied statistical analysis. Nine studies (28, 29, 31, 32, 34, 38, 40, 46, 47) applied regression models and bootstrapping methods to analyse the differences in costs and outcomes, whilst two studies (32, 46) adjusted for missing data. With trials being designed to assess the effectiveness of a new intervention for a specific population and CEAs/CUAs designed to account for benefits for even those not receiving a new health intervention, statistical analyses should be applied to address methodological challenges that may arise with the joint analysis.

### Economic evaluation findings and policy suggestions

Evidence of the findings of the economic evaluations of the caregiver interventions are summarised in Table 5. Of the 9 CEA studies and 5 CUA studies, four studies (31, 32, 38, 39) did not report outcomes using incremental cost effectiveness ratios (ICERs) due to small to no differences in the outcomes. Several of the studies reported the intervention as cost-effective (n=9) and cost-saving (n=5) especially when considering the long-term effects. A few of the studies found the intervention less likely to be cost effective. Charman et al (2021) found the predictive parenting intervention more costly and less cost-effective than its comparator (31). Similarly, Byford et al (2015) found the combination of pre-school autism communication therapy (PACT) and usual treatment to have higher costs and no significant difference in outcomes (32). Page et al (2016) found medication to be recommended for treatment of ADHD as it maximizes the societal cost compared to group behavioural therapy (42). With the willingness to pay threshold being arbitrary, most of the studies used varied thresholds to determine cost-effectiveness of the interventions.

**Table 5:** Summary of findings and policy implications.

| Study | Intervention(s) | Type of economic evaluation | Costs | Incremental cost effectiveness ratio (ICER) | Willingness-to-pay (WTP) threshold | Policy implications |
| --- | --- | --- | --- | --- | --- | --- |
| Scavenius et al (28) | CiC<sup>c</sup> is a manual-based program | Cost effectiveness analysis | Initial development costs \$91,916 (671,265 DKK)<br>Setup cost of training and running costs \$67,648 (494,034 DKK)<br>Cost per treated child \$1,178 (8,601 DKK) | \$1,707 (12,465 DKK) per effect size in PSOC<br>\$3,272 (23,892 DKK) per effect size in HSQ | \$4,117 per family for one effect size in PSOC score | CiC may be a cost-effective alternative for improving parental competence, compared to other BPT programs. |
| Sayal et al (29) | Parent only intervention<br>Combined parent-teacher intervention | Cost effectiveness analysis | Parent-only intervention £90<br>Combined interventions £107 | Parent-only intervention: £29 per one point | £31 per one-point improvement in the ADHD index | For children at risk of ADHD, an intervention programme for parents and teachers was |
|  |  |  | Incremental cost parent-only programme: £73<br>Incremental cost of the combined programme: £123 | improvement in the ADHD index<br>Combined intervention: £134 per one point improvement in the ADHD index |  | not associated with improvement in core ADHD symptoms.<br>The parent-only intervention programme suggested some potential for cost-effectiveness |
| Segal et al (30) | iBASIS–Video Interaction to Promote Positive Parenting (iBASIS-VIPP) intervention | Cost analysis and Cost-effectiveness analysis | Total cost of iBASIS-VIPP delivery (including TAU services): A \$5477 (US \$3850) per child | Cost per reduction in an ASD diagnosis at age 3 years: A \$37 181 (US \$26 138) | Not stated | iBASIS-VIPP is likely highly cost-effective.<br>iBASIS-VIPP likely represents a good-value societal investment. |
| | | | Total cost of TAU group: A \$346 (US \$243) per child<br>Cost difference: A \$5131 (US \$3607) per child | NPV cost savings: A \$10 695 (US \$7519) per child enrolled in iBASIS-VIPP (at 12 years age)<br>Return of investment: A \$3.08 (and US \$3.08) for each dollar invested in iBASIS-VIPP. | | Identifying pre-emptive interventions that are efficacious and represent good value is an important input to resource allocation decisions. |
| Sonuga-Barke et al (44) | New Forst Parenting Programme (NFPP) | Cost analysis (initially planned to do a cost utility analysis) | Total NFPP cost per family: £1591<br>Total IY cost per family: £2103<br>Cost difference: £512<br>NFPP intervention costs: £1081<br>IY intervention costs: £1569 | N/A | N/A | The finding that NFPP may be less costly than IY supports a revision of NICE's recommendations in favour of group rather than individual services.<br>Both individually delivered and group-based PT should be made available to families of children with preschool ADHD. |
| Charman et al<br>(31) | Predictive Parenting intervention | Cost effectiveness analysis | Predictive Parenting<br>Total NHS/PSS costs:<br>£789.77<br>Total public sector costs:<br>£1,393.42<br>Total societal costs:<br>£3,092.87<br>Psychoeducation<br>Total NHS/PSS costs:<br>£434.48 | No difference in QALYs | £30,000 per QALY gain | Predictive Parenting was more expensive than Psychoeducation, with a low probability of being more cost-effective. |
|  |  |  | Total public sector costs:<br>£700.19<br>Total societal costs:<br>£2,181.74 |  |  |  |
| Byford et al (32) | Pre-school Autism Communication therapy | Cost effectiveness analysis | Total cost PACT intervention: £4,105 per child<br>PACT + TAU<br>Total health, education and social service costs: £6,539 per child | Not explicitly stated | Varied | PACT is associated with significantly greater costs and no significant difference in outcome, and thus should not be recommended as a cost-effective in addition to TAU.<br>PACT plus TAU would only have a higher probability of |
|  |  |  | Total societal cost: £57919<br>TAU<br>Total health, education and social service costs: £2050 per child<br>Total societal cost: £56534 |  |  | being cost-effective compared to TAU alone if society is willing to pay extra for improvements in outcome. |
| Nystrand et al (33) | COPE Connect intervention.<br>Comet intervention<br>Incredible Years (IY) intervention | Cost utility analysis | Total cost per child<br>Comet: \$931<br>Connect: \$334<br>Incredible Years: \$1302<br>COPE: \$478 | Comet: \$972 per DALY averted<br>Connect: dominant | US\$ 15,000 per DALY | Parenting interventions are cost-effective in the longer run in comparison to a waitlist control. |
| | | | Bibliotherapy: \$14 | Incredible Years: \$1224 per DALY averted<br>COPE: dominant<br>Bibliotherapy: dominant | | Bibliotherapy or COPE are the most efficient options when comparing interventions to one another. Optimal decision for investment should be based on budget considerations and priority settings. |
| Tsiplova et al (34) | Parenting coaching intervention | Cost analysis | Public payer<br>PC group cost: \$6594 per family | N/A | N/A | The PACE Coaching project demonstrated that a parent coaching intervention can be |
| | | | ECT group cost: \$4079 per family<br>Mean incremental difference: \$2515<br>Societal<br>PC group cost: \$23,925<br>ECT group cost: \$16,931<br>Mean incremental difference: \$6994 | | | successfully implemented in a community setting.<br>The present cost analysis identified and measured major cost components of the coaching intervention which could be valuable for public funding and planning decisions to serve this population |
| Kuklinski et al (35) | First Step Next (FSN) intervention | Cost effectiveness analysis | Total cost of delivering FSN: US\$3,226 per student<br>Total cost of delivering combined intervention: US\$3,801 per student | ADHD FSN: \$12,433 per case reduction<br>Combined intervention: \$8,503 per case reduction<br>CD FSN: \$14,661 per case reduction | Varied | The combined intervention was more cost effective.<br>Improvement in comorbid ADHD and CD was the costliest to achieve, followed by CD, and then ADHD. |
| | | | | Combined intervention:<br>\$11,051 per case reduction<br>ADHD plus CD<br>FSN: \$16,909 per case reduction<br>Combined intervention:<br>\$12,131 per case reduction | | |
| Nystrand et al<br>(36) | Parent training programmes (COPE, Comet, Incredible Years, Connect) | Cost benefit analysis | Intervention costs per child<br>Comet: € 817<br>Connect: € 295<br>Incredible Years: € 1142<br>COPE: € 417<br>Self-help book: € 13 | Benefit cost ratio<br>Comet: € 7<br>Connect: € 10.61<br>Incredible Years: € 5.96<br>COPE: € 15.80<br>Self-help book: € 328.04 | Not stated | All the evaluated interventions within this study appear to be of good value-for money and yield substantial societal returns when adapted to various community populations, evidence that is relevant for local decision-making. |
| Tran et al (41) | Child Life and Attention Skills (CLAS) program and parent-focused treatment (PFT) | Cost effectiveness analysis | Total cost per patient<br>CLAS: \$1559<br>PFT: \$710<br>TAU: \$0<br>Incremental cost per patient<br>PFT: \$710<br>CLAS: \$1559<br>(compared to TAU) | CLAS versus TAU<br>\$3997 per disordered case<br>PFT versus TAU<br>\$3227 per disordered case<br>CLAS versus PFT<br>\$4994 per disordered case | Not stated | Nonpharmacological behavioural interventions can be viable, cost-effective treatment options for children with ADHD-I.<br>Future investigations are needed to evaluate cost-effectiveness of treatment for improving specific impairments associated with ADHD-I. |
| Page et al (42) | Group behavioral parent training | Cost analysis | Behavior modification (group parent training)<br>cost: \$961<br>Low dose of stimulant medication cost: \$1689 | N/A | N/A | Medication is more likely be recommended than behavioural treatment and used as a first line treatment for ADHD, despite its greater cost, and this approach maximizes the societal cost of ADHD treatments. |
| Penner et al (37) | Parent delivered Early Start Denver Model (ESDM) | Cost effectiveness analysis | Provincial perspective costs<br>Status Quo (SQ): \$186,000 per person | ESDM-I vs SQ: \$23,000/DFLY | Varied | Caregiver costs were a significant driver in cost-effectiveness estimates; consequently, from a societal |
| | | | ESDM-Parent Delivered model (ESDM-PD): \$178,000 per person<br>ESDM Intensive model (ESDM-I): \$199,000 per person | ESDM-I vs ESDM-PD: \$58,000/DFLY | | perspective the pre-diagnosis intensive ESDM generated both cost-savings and enhanced outcomes relative to both the status quo and pre-diagnosis parent-delivered ESDM. |
| Nystrand et al (38) | Parent management Training (PMT) programme | Cost effectiveness analysis | CPP intervention total cost per child: \$584 | Small difference in QALY gains | Varied | The results show that stacking treatments such as child-CBT and PMT may be money well spent compared<br>to PMT only.<br>Although costs are relatively small for the child component, investment in delivering both PMT and CPP depends on the willingness-to-pay for such a prioritisation. |
| Ferguson et al (43) | ABA- based parent training | Cost analysis | Total project cost: £1,509.3 per parent | N/A | N/A | A telehealth training platform should be considered by behaviour<br>analysts to expand their knowledge and increase the reach of their services, which could have benefits for parents who are not able to avail of training locally.<br>Additional cost savings identified highlight the potential of a telehealth platform for service provision. |
| O'Farrelly et al (39) | Video-feedback Intervention to promote Positive Parenting and Sensitive Discipline (VIPP-SD) | Cost effectiveness analysis; cost consequence analysis | 5 month follow up<br>VIPP-SD intervention total costs: £1827.55<br>Usual care total costs: £510.18<br>24 month follow up<br>VIPP-SD intervention total costs: £ 3131.93<br>Usual care total costs: £ 1525.38 | Very small differences in QALYs | > £800 for a 1-point improvement | VIPP-SD is more costly and more effective than usual care.<br>PPACS is not associated with a WTP threshold to support decision-making (compared with QALYs with a NICE WTP threshold of £20,000–30,000 per QALY), it is not possible to come to any firm conclusion about the relative<br>cost-effectiveness of VIPP-SD in the short term. |
| Gibbard et al (40) | Parent based intervention (PBI) | Cost effectiveness analysis | Cost per child<br>PBI: £96<br>General care: £80.83<br>Cost per parent<br>PBI: £31.80<br>General care: £5.78 | £10.97 per score gained | Not explicitly stated | The results of this study are supportive of the effectiveness of this parent intervention programme, employing the setting of specific linguistic targets within the child's naturalistic setting. This method of intervention also would not incur significant increases in cost per outcome gained over current practice and may, with modifications, result in cost savings. |
| Royston et al (46) | Stepping Stones Triple P (SSTP) programme | Cost effectiveness analysis (and cost utility analysis) | Cost per participant:<br>Intervention delivery £270<br>Training cost per participant £26 | Cost saving of –£1057.88 per participant<br>Very small differences in QALYs | £13,000, £20,000 and £30,000 per QALY gained | Stepping Stones Triple P is probably value for money to deliver, but decisions to roll this out as an alternative to existing parenting interventions or treatment as usual may be dependent on policymaker willingness to invest in early interventions to reduce behaviours that challenge. |
| Tinelli et al (48) | Preschool Autism Communication Trial (PACT) intervention | Cost consequence analysis | Total service perspective (health, education, and social services): England € 61,104; Ireland € 80,838; Italy € 31,204; Spain € 129,339<br>Total societal perspective (including parental productivity losses and | N/A | N/A | PACT is cost-saving over time from a societal perspective, even though we confirmed that, at 13 months post-delivery, PACT is more expensive than usual treatment (across all countries) when given to preschool autistic children. |
|  |  |  | informal care): England € 465,567; Ireland € 490,634; Italy € 366,299; Spain € 368,496. |  |  | After 6 years, we found that PACT has lower costs than usual treatment in terms of unpaid care provided by parents (in all countries). |
| Shimabukuro et al (47) | Well Parent Japan (WPJ) programme | Cost effectiveness analysis | | 6,707 (JPY) (\$ 61.93) per QALY gained | 10,000 JPY (\$ 108.30) and 20,000 JPY (\$ 216.60) | The cost-effectiveness analyses showed intervention costs to be modest and the programme cost-effective. |

### Methodological issues of economic evaluations

Although many of the included economic evaluations were rated average or good, many had methodological limitations, emphasising the challenges often faced in the conduct of economic evaluations of caregiver interventions for children with DDs. A few factors impact on the validity of conclusions drawn in economic evaluation such as accuracy of cost information, choice of discount rate, scope, modelling and time horizon and perspective. However, three main methodological challenges were identified from the literature as representative of the methodological issues more specific to conducting economic evaluations of caregiver interventions for young populations: 1) measuring and valuing cost, 2) measuring health outcomes and 3) time horizon analysis.

### Measuring and valuing costs

A prominent challenge related to cost measurement and valuation was the cost categories included. The studies identified exclusion of important cost inputs that would over-or underestimate the cost-effectiveness outcome. For societal costs, Scavenius et al (2020) did not account for productivity loss costs (28), and Nystrand et al (2019) did not include caregiver health and well-being costs and the costs of behaviour problem consequences in adulthood which would be significant cost offsets in the long term (33). Regarding provider costs, Tsiplova et al (2022) excluded certain costs such as intervention development costs, and training costs (materials, coaches travel expenses, coach training) (34), and Nystrand et al (2020) included only medication costs in the healthcare resource use (36). The accuracy of resources measurement was potentially limited by self-reporting of resource use for instance informal care and parent time costs in two studies (32, 41). Another challenge noted in two studies (35, 41) was the lack of data, which limited the costs included in the analysis. Further, the selected perspective of the analysis was considered narrow in one study (33), because mental health interventions may have a broader societal impact, and interact with other sectors such as education, social services, justice, and voluntary sectors. Notably, Penner et al (2015) found that caregiver costs were a significant cost driver in the cost-effectiveness estimates further highlighting the importance of accounting for all relevant costs (37).

### Measuring health outcomes

Measuring health related quality of life in children remains contentious because of the descriptive system or valuation perspective. Sayal et al (2016) utilized two measures CHU 9D and EQ5D-Y which showed slight variance in results likely from the differences in the valuation processes employed (29). One study (33) used parental proxy rather than self-reported outcomes, which may be associated with misreporting. Also, based on the other health outcome measures applied in two studies (32, 39) it was noted that QALYs measures have mostly been applied in adult populations and may not be representative of the younger populations. Further, utility values were not available for some countries (36), or were condition-specific for instance per case reduction (37, 41), which limits the use of QALYs as outcome measures. Regarding application of DALYs, Nystrand et al (2019) stated this health outcome was limited by availability of a single disease weight for conduct disorders making it difficult to model changes in disorder severity without significant assumptions about the distribution of disorder and disability weight by severity. Moreover, children with some levels of problems who would benefit from the intervention and the benefits incurred by parents, siblings and teachers were excluded, which would underestimate the health gains attributable to the interventions (33).

### Time horizon

Caregiver interventions for developmental disorders may result in long-term benefits and the choice of time horizon analysis should aim to capture all meaningful costs and outcomes. One trial based economic evaluation studies (41), modelled shorter timeframes due to lack of published data. However, the intervention showed a sustained effected with indication of it being even more cost-effective in later years of life. Similarly, O’Farrelly et al (2021) found that the short time horizon was a limitation as the participants were less likely to incur costs for service use relating to behavioural problems during the preschool period but instead when the child is older (39). Two studies (37, 39) reported that the use of multiple data sources to extrapolate results over longer periods and to different population should be used with caution because this would require many assumptions and huge uncertainties which would likely produce poor estimates of the results.

## Discussion

This review identified economic evaluations of caregiver interventions for children with DDs and synthesized evidence on the methods adopted and their associated challenges. Thirty studies were identified that carried out both partial and full economic evaluations. Most of the studies evaluated interventions delivered through group programmes (47%) compared to individual based platforms (41%). One striking feature of this review is the heterogeneity of the economic evaluation approaches presented. The majority of studies (70%) were trial based economic evaluations, which are considered suitable for inferring cost-effectiveness for interventions with no prior evidence (49). Current trial based economic evaluations have limited time horizons, often less than one year. Extending the time horizon of economic evaluations may lead to more favourable estimates (50), especially for interventions with long-term effects, as in the case of caregiver interventions. In our review seven studies extrapolated data over a one-year time horizon. However, extrapolation of data for long term models should be applied cautiously to minimize errors from multiple sources of data and the related assumptions. The uncertainties of parameters contribute to the overall model uncertainties whose potential impact can be assessed through scenario and sensitivity analyses, which are important considerations for economic evaluations (19). Modelling studies may help address some of the issues of RCTs with longer period projections of the cost estimates and outcomes, and should use available evidence of real-world data and assumptions. Our review identified only four studies, which applied a decision analytical modelling approach such as decision trees or Markov models. Within any economic evaluation modelling, there is a balance between adequately simplifying complex health states and interventions to allow model parameterisation, while avoiding oversimplification to the extent that findings are not representative of the real-world context (19). Even with diverseness of the methods applied, more than half (64%) of the reviewed studies found that the caregiver interventions were cost saving or cost-effective, while improving quality of life for the children and parents.

The studies reported varied levels of details of cost components used in the economic evaluations, with most studies including the major components as start-up and implementation costs. A few (18%) of the reviewed studies included costs from other sectors or cost offsets as important cross cutting and relevant costs which significantly contribute to wholistic care of children with DDs. Children with DDs require various services often extending beyond healthcare, for instance social services, education, rehabilitation, and criminal and justice systems. Suhrcke et al (2008) conducted a systematic review demonstrating that for child and adolescent mental illness, only 6% of costs accounted for healthcare, with majority of the costs in other sectors (i.e. social services, education, productivity) (51). Moreover, the service needs for children with DDs changes with the setting and age during their lifetime (21). Therefore, quantifying service utilization for an economic evaluation of caregiver interventions may be challenging. Some of the reviewed studies excluded certain costs due to lack of data. For instance, one study (41) was unable to assess all costs necessary for a full societal approach, another study (33) applied a narrow costing perspective, which likely missed different social impacts, one other studies (35) used a limited health system perspective and only cost offsets for one episode of care. While inclusion of such costs may increase the intervention cost, the incremental costs of the intervention may also reduce. Adopting a narrower costing perspective may fail to incorporate other relevant sectors, which may bias study findings when most service utilization is beyond the healthcare system (1). The economic benefits that accrued over time are realized not only by the participant and family involved, but also by other sectors and society in general (40). In addition, the impact on productivity for caregivers may be considered in terms of time spent at work, and more often primary caregivers of children with DDs reduce working hours, give up work or change jobs to provide care (1). Two main challenges with quantifying and valuing productivity are differentiating hours of care provided related to the DD compared to usual care and measuring productivity for children for their future earnings (52). One of the reviewed studies (28) excluded productivity costs of parents and indicated possible underestimation, whereas two studies (33, 41) included parent time costs which added value to the findings. Further, Penner et al (2015) indicated caregiver costs as a significant cost driver for the intervention (37). Although including productivity costs requires careful consideration, failure to capture productivity costs may potentially underestimate the resources use and effectiveness of the intervention (1).

Most of the included studies used cost effectiveness methods. Although informative, CEAs measure condition specific outcomes that are not directly comparable to other interventions targeting the same related problems or those across different diagnostic areas. In addition, the use of clinical measures undermines the likelihood of finding improvements that may be relevant to general well-being and everyday life, for instance improvement in quality of life. This is a key aspect of caregiver interventions that may have an impact on different areas of the children’s and parents’ lives. Therefore, cost utility studies which measure health outcomes with a generic health status that considers both mortality and the quality of life with morbidity would be preferred. In this review, only eight studies conducted CUAs with QALYs, DALYs and DFLYs used as the outcome measures. Measuring effectiveness of DDs is challenging and involves assessment of physical, behavioural, psychological, cognitive and social domains (1). In this regard, our review highlighted methodological challenges towards measuring QALYs and DALYs in mental health and for the younger population.

First, the use of DALYs as an outcome may be limited by data availability on disease weights for specific DDs and the severity levels. For instance, Sampaio et al (2015) noted that conduct disorder (CD) doesn’t have differential severity weights, and significant assumptions on the disorder distribution and disability weights would be required to model changes in disorder severity (53). Consequently, children with some levels of CD who would benefit from the intervention were excluded from the analysis, which would likely underestimate the health gains attributed to the reduction in disorder severity (53). A better approach would specify weights based on disorder severity as a reflection of heterogeneity in health (54). Second, the use of QALYs and the maximization approach to resource allocation in relation to mental health remains a contentious and understudied area (55). Specifically, QALY measures may be “mis-valued”, insensitive and not specific to all mental health issues. Nonetheless, in failing to use QALYs there is risk of marginalization of mental health interventions in priority setting and budgeting processes.

Third, there are challenges in methods of measuring and valuing health related quality of life in children, which have been studied to a limited extent than the adult population. In a recent review, some of the challenges highlighted included how to elicit informed preferences, the generation and use of combined adult and adolescent preferences and the appropriateness and acceptability of valuation tasks for children and adolescents (56). Comparably, one of the included studies (32) was limited to measures available for children seven years of age and above, which was not suitable to the study population of pre-school children. Mihalopolous et al (2015) highlighted that the study was limited by the availability of adult population parameters which would not be representative of anxiety problems and the disease duration estimates in young children (57). Thus, the use of different health related quality of life measures may show slight differences in the results, like one of the included studies (29). Although the indirect approaches of utility measurement from multi-attribute utility instruments including EuroQol 5 dimensions (EQ-5D) and Health Utilities Index (HUI) are more appropriate to measure health status of children with DDs, deriving the parameter estimates can be challenging because of age-appropriateness, the domains, and methods to derive utilities (58). Despite the challenges, there remains extensive opportunity for improvement of these estimates. Fourth, the application of utility values from other countries may be less accurate than country specific as highlighted in one of the reviewed studies (38). In addition, there is limited data on utility values for some DDs such as autism spectrum disorder (ASD) as noted in one of the reviewed studies (37). The valuation of child and adolescent outcome measures remains a challenging research area that requires further empirical evidence to inform best practice.

### Strengths and limitations

A strength of this review is the application of the Arksey and O’Malley framework for scoping reviews which encourages collaborative engagement in the review process (22). This involved identifying and developing the overarching research question, and the key terms to identify relevant studies. A notable limitation of this review was the relatively few studies to review with a strong focus on caregiver interventions for children with NDDs which may restrict generalizability of the findings while highlighting important areas for future research. While the search strategy was specific to NDDs, most papers focused on developmental disabilities all in high-income countries, evidently showing a literature gap. Costing and cost effectiveness data is highly context specific considering contextual factors that are important such as out of pocket costs, different health systems and different quality of life perspectives amongst others. This provides directions for future research across different contexts for example the African region.

### Conclusion

This review mapped economic evaluations of caregiver interventions for children with DDs and highlighted the gap in evidence and methodological challenges. Whilst economic evaluation analyses in this area are scarce, emerging data from common DDs was promising in the quest for cost saving and cost-effective interventions that improve quality of life of both the child and parent/caregiver. Caregiver interventions are a promising avenue to strengthen access and reduce costs associated with health services for children with DDs. This review has provided an overview of evidence on DDs which is a growing priority across many areas of paediatrics. Future research should consider the development of appropriate outcome measures and measurement of all relevant costs for this heterogenous population. Prioritizing more economic evaluation studies in this area would inform decision-making on efficient resource allocation, promote inclusivity and equitable access of services for children with DDs within the health system.

## Data Availability

All data has been included in the article and additional supporting information files.

## Acknowledgements

CH receives support from the National Institute for Health and Care Research (NIHR) through the NIHR Global Health Research Group on Homelessness and Mental Health in Africa (NIHR134325) using UK aid from the UK Government. CH also receives support from Wellcome grants 222154/Z20/Z and 223615/Z/21/Z.

For the purpose of open access, the authors have applied a Creative Commons Attribution (CC BY) licence to any Author Accepted Author Manuscript version arising from this submission.

## Supporting information

S1 Appendix: Search concepts and the corresponding key words used (concepts were combined using the Boolean operator “AND”)

S2 Appendix: CHEERS 2022 Checklist AND Gates Reference Case Principles

S3 Appendix: Quality assessment of economic evaluations using the Drummond checklist

